# Individual treatment effect estimation in the presence of unobserved confounding using proxies: a cohort study in stage III non-small cell lung cancer

**DOI:** 10.1101/2021.10.30.21265597

**Authors:** Wouter A.C. van Amsterdam, Joost. J.C. Verhoeff, Netanja I. Harlianto, Gijs A. Bartholomeus, Aahlad Manas Puli, Pim A. de Jong, Tim Leiner, Anne S.R. van Lindert, Marinus J.C. Eijkemans, Rajesh Ranganath

## Abstract

Randomized Controlled Trials (RCT) are the gold standard for estimating treatment effects but some important situations in cancer care require treatment effect estimates from observational data. We developed “Proxy based individual treatment effect modeling in cancer” (PROTECT) to estimate treatment effects from observational data when there are unobserved confounders, but proxy measurements of these confounders exist. We identified an unobserved confounder in observational cancer research: *overall fitness*. Proxy measurements of overall fitness exist like performance score, but the fitness as observed by the treating physician is unavailable for research. PROTECT reconstructs the distribution of the unobserved confounder based on these proxy measurements to estimate the treatment effect. PROTECT was applied to an observational cohort of 504 stage III non-small cell lung cancer (NSCLC) patients, treated with concurrent chemoradiation or sequential chemoradiation. Whereas conventional confounding adjustment methods seemed to overestimate the treatment effect, PROTECT provided credible treatment effect estimates.

## Introduction

Randomized controlled trials (RCT) are the gold standard for estimating treatment effects but there are important situations in cancer care where treatment effect estimates from observational data are needed. First, study participants of cancer RCTs are generally younger and in better overall health when compared to the real-world population *(1–3)*. Therefore, RCTs provide no direct evidence for applying the treatments in older and weaker patients, as these parts of the population are not covered by the trials. The effect of a treatment in these subpopulations could be estimated in observational data to investigate whether a RCT with extended inclusion criteria is indicated. Second, there is a constant research effort to discover new predictive biomarkers. Predictive biomarkers reveal parts of the biological behavior of the tumor that are related to the treatment effect and can be used to select the optimal treatment for a patient *(4)*. Before RCTs with new predictive biomarkers can be conducted, preliminary evidence on their association with the treatment effect is needed based on observational data. Finally, there are situations where RCTs are possible but may be deemed unethical, such as in pediatric oncology or end-of-life care. In these situations, observational data may provide the only basis to inform treatment policies.

Estimating treatment effects in observational data requires knowing what the confounders are of the treatment – outcome relationship. In cancer care, the most effective treatment is often also the most intensive treatment. The *overall fitness* of a patient determines what treatment intensity they can endure. Therefore, overall fitness is the central confounder. The treating physician will form an implicit assessment of overall fitness that is partly based on the subjective impression of a patient. As there is no record of this implicit assessment, traditional confounding adjustment methods cannot be used. However, proxy measurements of fitness are available, such as performance score. We developed a method named ‘Proxy based individual treatment effect modeling in cancer’ (PROTECT). PROTECT uses proxy measurements of fitness to reconstruct the distribution of the unobserved confounder and to adjust the treatment effect with this reconstructed confounder. In addition to modeling the confounder, PROTECT allows for incorporation of biomarkers of the biological behavior of the tumor. These biomarkers together with the patient overall fitness are used to predict the individual treatment effect.

We apply PROTECT to stage III Non-Small Cell Lung Cancer (NSCLC). Newly diagnosed patients with stage III NSCLC have two curative treatment options: concurrent chemotherapy and radiotherapy, or sequential treatment with chemotherapy followed by radiotherapy *(5)*. According to the meta-analysis of RCTs by Aupèrin et al., concurrent treatment leads to better overall survival (hazard ratio, 0.84; 95% confidence interval, 0.74 to 0.95) *(6)*. Concurrent treatment is more intensive and has a higher risk of severe toxicity *(6)*. Treatment guidelines recommend to give sequential treatment to patients with lower overall fitness *(5, 7)*. These patients are more likely to experience treatment toxicity under concurrent chemoradiation that would require negative adjustment or cessation of the treatment. This suggests that the survival benefit of concurrent treatment is absent or reversed in patients who are in lower overall fitness. As the real-world population contains older and weaker patients than the RCTs, an important question is whether the average treatment effect estimate from the RCTs is valid in the real-world population, and whether this average treatment effect applies to all patient subgroups.

In this study we present PROTECT as a method for estimating both the average treatment effect and individual treatment effects in real-world cancer populations from observational data. The method is applied to a multi-center observational cohort of stage III NSCLC patients, comparing concurrent chemoradiation with sequential chemoradiation.

## Results

### PROTECT

The objective of PROTECT is to estimate treatment effects from observational data. This requires knowing what the confounders are of the treatment – outcome relationship. In multiple discussion rounds with experts in oncology, thoracic oncology, radiotherapy, radiology, causal inference, statistics and epidemiology, we identified the confounders of the treatment – outcome relationship in cancer.

Patients who are in good overall health will more often be prescribed the most intensive and effective treatments as they can tolerate these treatments better than patients with lower overall fitness. Patients with better overall fitness also have better outcomes regardless of their treatment. This means that the overall fitness of a patient is a confounder of the treatment – outcome relationship. A treating physician will form an implicit assessment of the overall fitness of a patient that is partly based on a subjective impression of the patient. As there is no record of this implicit assessment of fitness, for the purpose of research this confounder is unobserved. Only the patient characteristics like performance score and age are available.

In addition to the overall fitness of a patient and the treatment they receive, an important factor for the variation in patient outcomes is the biological behavior of the tumor. There are different biomarkers of tumor behavior that are known in the clinical process, like histologic subtype or intra-tumor genetic heterogeneity *(8)*. These biomarkers are related to prognosis and / or the treatment effect. Tumor behavior is fundamentally unobservable in the sense that neither the physician nor the researcher observe this behavior fully. Only the biomarkers are available.

Patient fitness and tumor behavior are thus two unobserved variables that induce relationships between the observed patient characteristics, the biomarkers, the treatment decision and the outcome. The causal relationships between these variables are represented in a causal directed acyclic graph (DAG), shown in Figure 1. For different cancer settings, different markers of patient fitness and tumor behavior may be relevant. Filling in application specific variables in the DAG is the first step of the PROTECT method. It could be that multiple choices for sets of variables are possible for a specific application. As explained in the appendix (section methods, estimation in a marginalized DAG), the PROTECT average treatment effect estimate is insensitive to the specific choice of variables.

**Figure 1.**
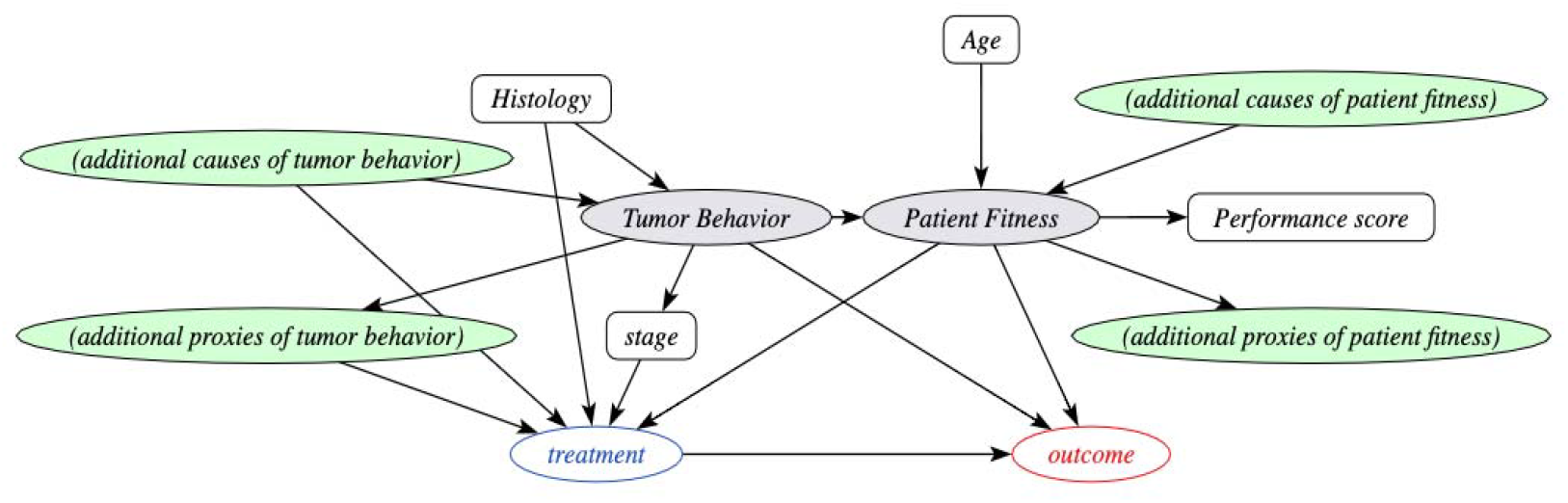
The behavior-fitness causal Directed Acyclic Graph (DAG) scaffold for cancer treatment decisions. Circles indicate variables, grey-shaded variables are unobserved. Arrows point from a cause variable to an effect variable. Tumor behavior and patient fitness are unobserved variables that induce correlations between the observed variables. The definition of the treatment variable and potentially the outcome variable vary per cancer setting. Depending on the specific situation, relevant additional cause variables and effect variables for tumor behavior and patient fitness should be selected. Estimating the effect of the treatment on the outcome (potentially conditional on the other variables in the DAG) is the target application of PROTECT. The presence of the unobserved confounder fitness implies that conventional confounding adjustment methods cannot be used to estimate treatment effects from observational data, whereas the proposed method PROTECT can. Filling in additional proxies and causes of tumor behavior and patient fitness in this DAG is the first step of PROTECT. PROTECT: proxy based individual treatment effect modeling in cancer.

### From the DAG to treatment effect estimation

Having established a DAG for observational cancer research, the question is if and how the treatment effect can be estimated from observational data. To answer this, we use Pearl’s Structural Causal Models framework *(9)*. The presence of the unobserved confounder overall fitness implies estimates of the treatment effect by direct conditioning on the observed variables (e.g. through multivariable regression or propensity score-based reweighting) would be incorrect as the back-door criterion is not fulfilled *(9–13)*. This does not rule out the possibility to estimate the treatment effect. Several methods exist for estimating treatment effects when there are proxy variables of unobserved confounders. Proxy variables are variables that are caused by the confounder, but do not causally influence the treatment decision and the outcome. Performance score is an example of a proxy variable for the confounder overall fitness, as performance score depends on fitness but does not cause overall survival or the treatment decision directly. One class of proxy methods relies on bridge functions *(12, 14–17)*. These methods leverage information from proxy measurements to reconstruct a bridge function that is sufficient for treatment effect estimation. To know if such a function can be estimated from the observed variables, these methods require additional assumptions. A frequently required assumption is that all variables are discrete *(12, 14, 17)*, which is an unnatural assumption for the confounder overall fitness, or that all variables are continuous *(12, 14)*, which is rarely the case in medical research. More flexible bridge function methods exist but these methods require complicated estimation procedures that require large sample sizes, which makes these methods unsuitable for many medical applications *(15)*.

An alternative approach is by estimating the joint distribution of the observed variables and the unobserved variables, using only the observed variables *(18)*. When sufficient information on the data generating process is available, this joint distribution can be estimated by modeling the data generative process directly. With this approach, each variable in the DAG is associated with an explicit structural equation that depends on the direct cause variables of this variable and random noise. If the joint distribution can be estimated, the treatment effect can still be calculated because the back-door adjustment formula can be applied using the estimated distributions of the outcome given treatment and fitness, and fitness given the cause variables and proxy variables of fitness *(9, 18)*.

Both proxy-based approaches require assumptions in addition to the DAG. These assumptions should be based on background knowledge. Background knowledge naturally comes in the form of parts of the data generating process. Modeling the data generating process directly thus makes formulating the right assumptions easier for clinicians and researchers. Moreover, it makes it more accessible for readers to assess the validity of the made assumptions. One example of such an assumption is the statement that performance score should be better for patients with higher overall fitness. This assumption can be expressed as a monotonicity restriction in the structural equation for performance score and helps estimating the joint distribution of observed and unobserved variables and thus the treatment effect. In PROTECT, the joint distribution is estimated by specifying parametric forms for all the structural equations. If the parameters for the structural equations can be uniquely estimated from the observed data, the treatment effect can be estimated despite the unobserved confounder (see also appendix, section methods, treatment effect estimation). Translating background knowledge into specified parametric distributions for the observed and unobserved variables in the DAG thus forms the second step of PROTECT.

### Model selection

It is possible that multiple choices for distributions are compatible with the available background knowledge for a specific research question. To reduce dependence of the treatment effect estimate on the specific choices for distributions, we introduce a data-driven model selection procedure. The model selection procedure is motivated by the fact that in the DAG, the unobserved variable overall fitness induces correlations between the proxy variables, treatment and outcome. The procedure uses cross-validation to verify whether these correlations are present in an estimated model by comparing the model predictions for one of the effect variables of fitness (proxies, treatment and outcome) with regression models based on only the direct causes of this variable, excluding fitness. If multiple models are selected in this step, they can be combined using a Bayesian model average. The appendix (section methods, model selection) contains a detailed report of this model selection procedure. After selection and estimation of the final model, the individual treatment effect is calculated as the difference in the expected outcome under the different treatments for each patient, conditional on their observed pre-treatment characteristics.

### Computation

Once the parametric distributions are fully specified, the posterior distribution over the parameters for the structural equations can be estimated using Markov chain Monte Carlo (MCMC) sampling. We implemented PROTECT using state-of-the art inference techniques. Specifically we employ the No-U-Turn Hamiltonian Monte Carlo *(19)* implementation from the NumPyro package *(20)* as NumPyro has JAX *(21)* as a back-end, enabling parallelized GPU-accelerated MCMC sampling. The code that implements PROTECT will be made freely available at a public online repository.

### Application to stage III NSCLC

We now apply PROTECT to stage III NSCLC patients to estimate the relative effect of concurrent chemoradiation versus sequential chemoradiation on overall survival measured from the day of the treatment decision.

### PROTECT step 1: definition of proxy variables and cause variables

In multiple discussion rounds with experts in pulmonary oncology, radiation oncology and radiology, the relevant proxy variables and cause variables for stage III NSCLC were selected. The additional variables included in the stage III NSCLC DAG are weight loss and estimated glomerular filtration rate (eGFR). Weight loss is an important proxy of tumor behavior, as patients with aggressive tumors tend to lose more weight due to the high disease burden. Renal function is an additional proxy variable for overall fitness in stage III NSCLC. The DAG is presented in Figure 2.

**Figure 2.**
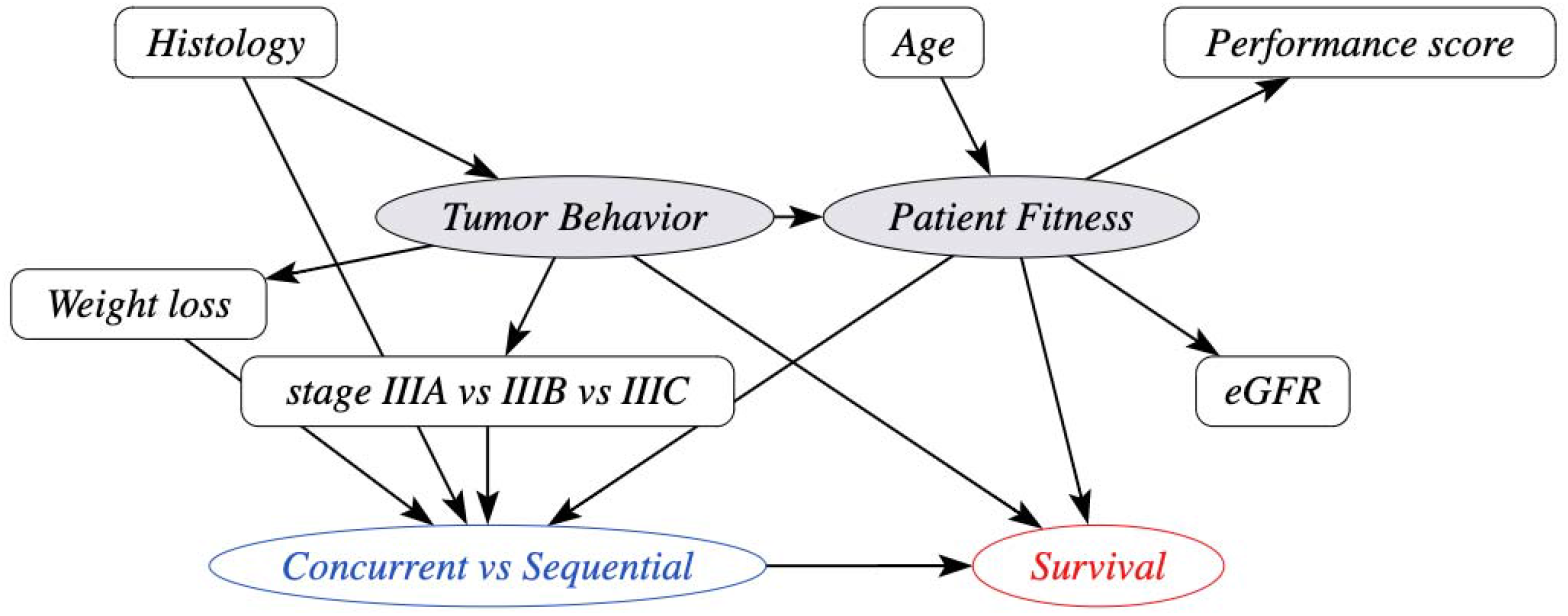
Causal Directed Acyclic Graph (DAG) with the variables involved in the treatment selection process and overall survival for stage III non-small cell lung cancer patients. Circles indicate variables, shaded variables are unobserved. Arrows point from a cause variable to an effect variable. This DAG is a direct extension of the behavior-fitness DAG scaffold from the PROTECT method in Figure 1. PROTECT: proxy based individual treatment effect modeling in cancer; eGFR: estimated glomerular filtration rate.

### PROTECT step 2 and 3: specification of distributions and model selection

We parameterized the joint distribution of observed variables and unobserved variables by specifying linear models for all structural equations indicated by the DAG, with link functions and error distributions as appropriate (i.e. linear regression for continuous variables, logistic regression binary variables, a linear proportional hazards model for survival *(22)*). To estimate individual treatment effects, the model for survival was augmented with treatment – covariate interaction terms. Details on the exact formulation of the model variants entered in the model selection procedure are presented in the appendix (section methods, pre-processing, parametric models and priors).

### Study Population

We identified 844 patients with 864 episodes of stage III NSCLC in 9 hospitals in the Utrecht region, the Netherlands, treated between 2009 and 2018. A total of 360 episodes were excluded based on the following exclusion criteria (more than one criterion can apply to an episode): the primary treatment plan had palliative intent (N=140), the primary treatment plan included surgery (N=49), the presence of a concurrent other tumor, including a second NSCLC (N=35), local re-irradiation for the recurrence of an earlier episode (N=9), Pancoast tumor (N=9), having a second episode of stage III NSCLC (N=8, only the second episode was excluded from the analysis), receiving radiotherapy in a different hospital (N=3), chemotherapy and radiotherapy in reversed order to prevent spinal canal invasion (N=3), emigration (N=1). Finally, 123 patients were excluded due to a missing value for weight loss.

The mean age of the 504 included patients was 64.9 (range 37 - 86), of which 300 (59.5%) were male. Substage IIIA accounted for 268 of the cases (53%). We observed 141 deaths in 224 patients who underwent concurrent chemoradiation (632 patient years) and 214 deaths in 280 patients who underwent sequential chemoradiation (603 patient years). Compared with the study population from the meta-analysis of RCTs *(6)*, our patients were older (median, 61.7 vs 66) and had worse performance scores (ECOG of 2 or greater: 1% vs 10%). In the appendix (section results, supplemental tables) a table with an extensive comparison is presented. The start of follow-up was imputed for 12.3% of the patients. The median survival time was 1.87 years, the median follow-up time for patients who were censored was 3.80 years. The last date of follow-up was February 6^th^, 2020. Patient characteristics are summarized in Table 1.

**Table 1.**
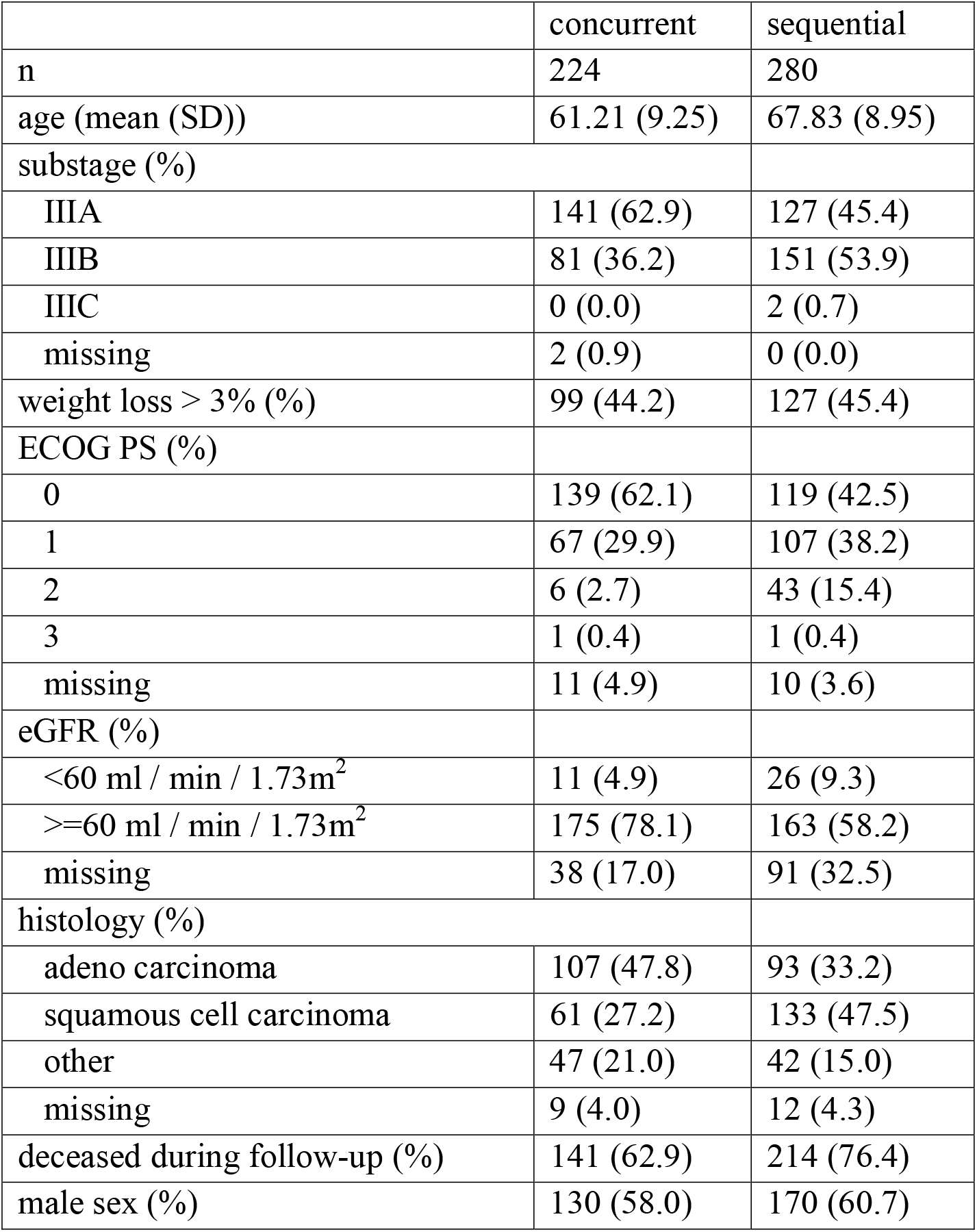
Baseline characteristics stratified by chemoradiation type: concurrent chemoradiation or sequential chemoradiation. Weight loss is defined as weight loss over 3% of the original weight over the six months preceding the start of follow-up. ECOG PS: Eastern Cooperative Oncology Group performance score; SD: standard deviation; eGFR: estimated glomerular filtration rate.

|  | concurrent | sequential |
| --- | --- | --- |
| n | 224 | 280 |
| age (mean (SD)) | 61.21 (9.25) | 67.83 (8.95) |
| substage (%) |  |  |
| IIIA | 141 (62.9) | 127 (45.4) |
| IIIB | 81 (36.2) | 151 (53.9) |
| IIIC | 0 (0.0) | 2 (0.7) |
| missing | 2 (0.9) | 0 (0.0) |
| weight loss > 3% (%) | 99 (44.2) | 127 (45.4) |
| ECOG PS (%) |  |  |
| 0 | 139 (62.1) | 119 (42.5) |
| 1 | 67 (29.9) | 107 (38.2) |
| 2 | 6 (2.7) | 43 (15.4) |
| 3 | 1 (0.4) | 1 (0.4) |
| missing | 11 (4.9) | 10 (3.6) |
| eGFR (%) |  |  |
| <60 ml / min / 1.73m<sup>2</sup> | 11 (4.9) | 26 (9.3) |
| ≥60 ml / min / 1.73m<sup>2</sup> | 175 (78.1) | 163 (58.2) |
| missing | 38 (17.0) | 91 (32.5) |
| histology (%) |  |  |
| adeno carcinoma | 107 (47.8) | 93 (33.2) |
| squamous cell carcinoma | 61 (27.2) | 133 (47.5) |
| other | 47 (21.0) | 42 (15.0) |
| missing | 9 (4.0) | 12 (4.3) |
| deceased during follow-up (%) | 141 (62.9) | 214 (76.4) |
| male sex (%) | 130 (58.0) | 170 (60.7) |

### Treatment Effect Estimation

Overall survival was significantly better for patients with concurrent chemoradiation compared to sequential chemoradiation (hazard ratio, 0.66; 95% confidence interval, 0.53 to 0.82). When estimating the treatment effect with multivariable cox-regression, adjusting for age, histology, weight loss, clinical substage, performance score and eGFR, concurrent treatment had a favorable survival (hazard ratio, 0.81; 95% confidence interval, 0.60 to 1.09). This treatment effect estimate is more extreme than the effect reported in the meta-analysis of RCTs (6) and possibly affected by residual confounding bias. In contrast, the average treatment effect estimated using PROTECT showed no benefit of concurrent over sequential treatment on average in our population (hazard ratio, 1.01; 95% credible interval, 0.68 to 1.53). An overview of the treatment effects is presented in Figure 3.

**Figure 3.**
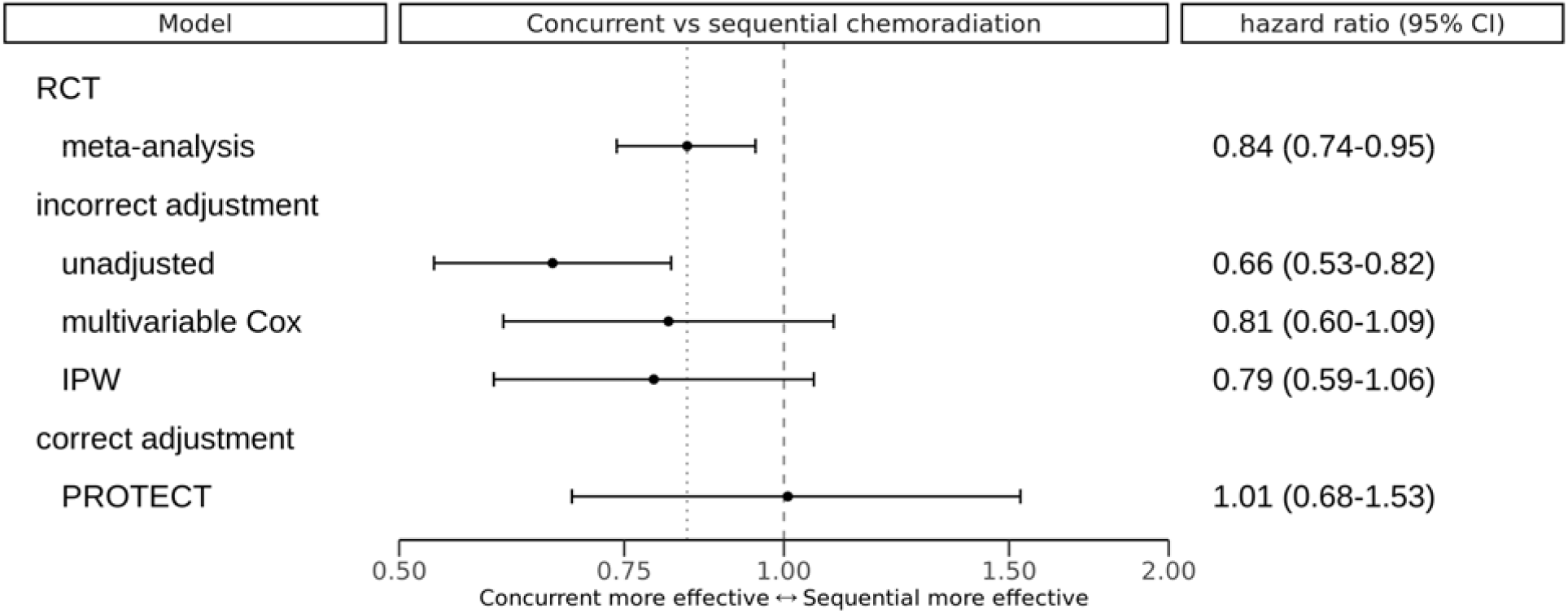
Overview of treatment effects estimated with different methods. The dashed vertical reference line indicates the null effect (hazard ratio of 1), the dotted reference line indicates the point estimate of the meta-analysis of RCTs by Aupérin et al. (6). IPW: inverse-probability of treatment weighted Cox-proportional hazards model. PROTECT: proxy based individual treatment effect modeling in cancer; CI: confidence interval, for PROTECT: credible interval; RCT: randomized controlled trial.

### Treatment Effect Modification and Individual Treatment Recommendations

According to the PROTECT individual treatment effect model estimate, the following variables were associated with a reduced effectiveness of concurrent treatment: clinical substage IIIB and IIIC, the presence of weight loss and adenocarcinoma histologic subtype. Age, ECOG performance score and eGFR were not related to treatment efficacy. Treatment effect modifications are presented in Figure 4.

**Figure 4.**
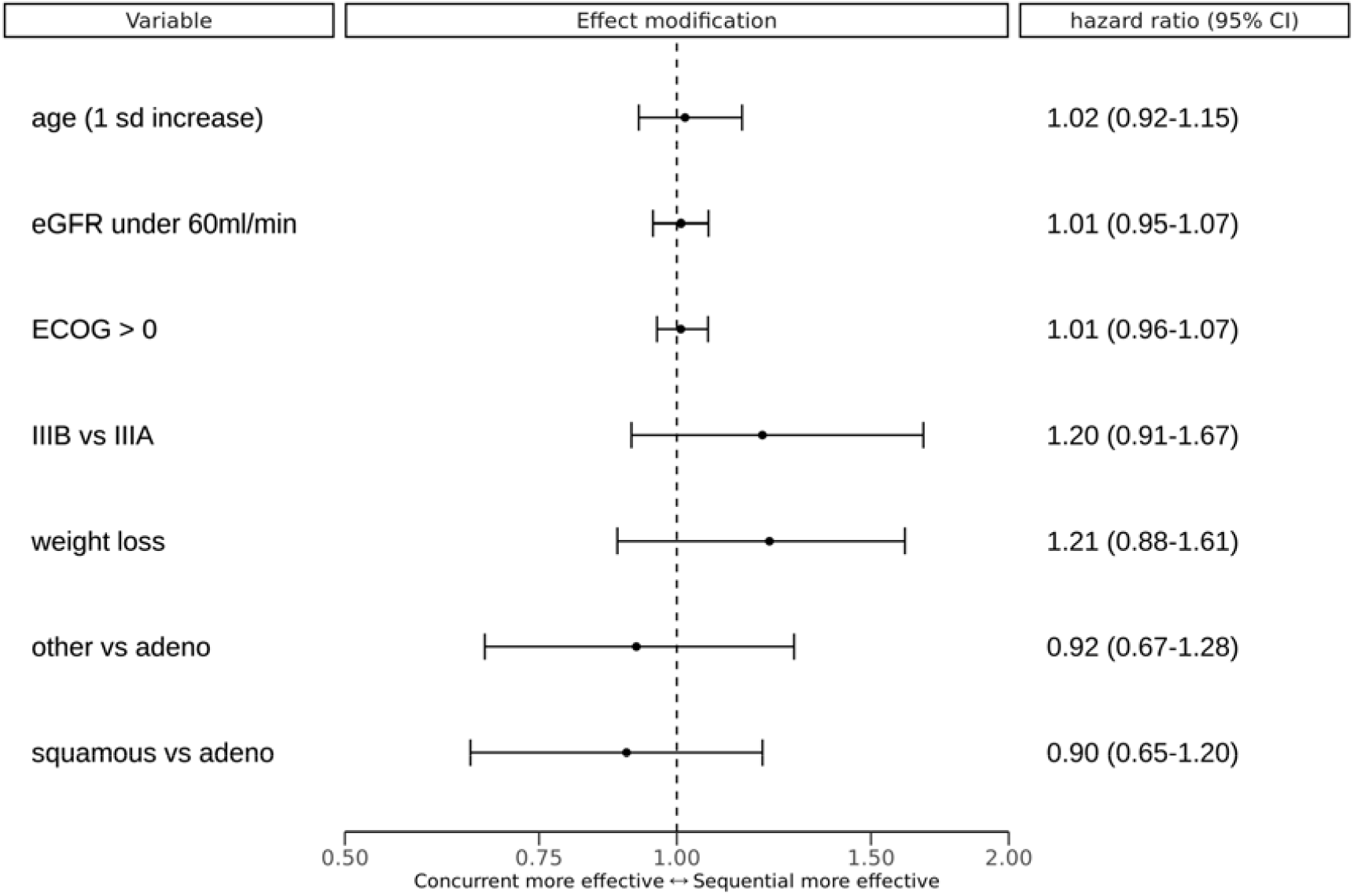
Differences in estimated treatment effect compared to the average treatment effect for a one unit increase per variable. A unit increase means switching from ‘no’ to ‘yes’ for binary variables, and a 1 standard deviation increase from the mean for continuous variables (age). These are step-function versions of the partial dependence functions as described by Friedman (23). ‘other vs adeno’ indicates the effect modification of other histology type compared to adenocarcinoma. ‘squamous vs adeno’ indicates the effect modification of squamous cell carcinoma compared to adenocarcinoma. CI: credible interval, eGFR: estimated glomerular filtration rate, ECOG: Eastern Cooperative Oncology Group performance score, IIIB: clinical stage IIIB or IIIC, IIIA: clinical stage IIIA, Weight loss is defined as weight loss over 3% of the original weight over the six months preceding the start of follow-up.

For each patient PROTECT predicted the probability that concurrent treatment would lead to improved survival compared with sequential treatment, based on the pre-treatment variables. For 274 out of 504 patients (54.4%) this probability was greater than 50%. See Figure 5 for an overview of the predicted treatment benefit expressed as a hazard ratio per patient.

**Figure 5.**
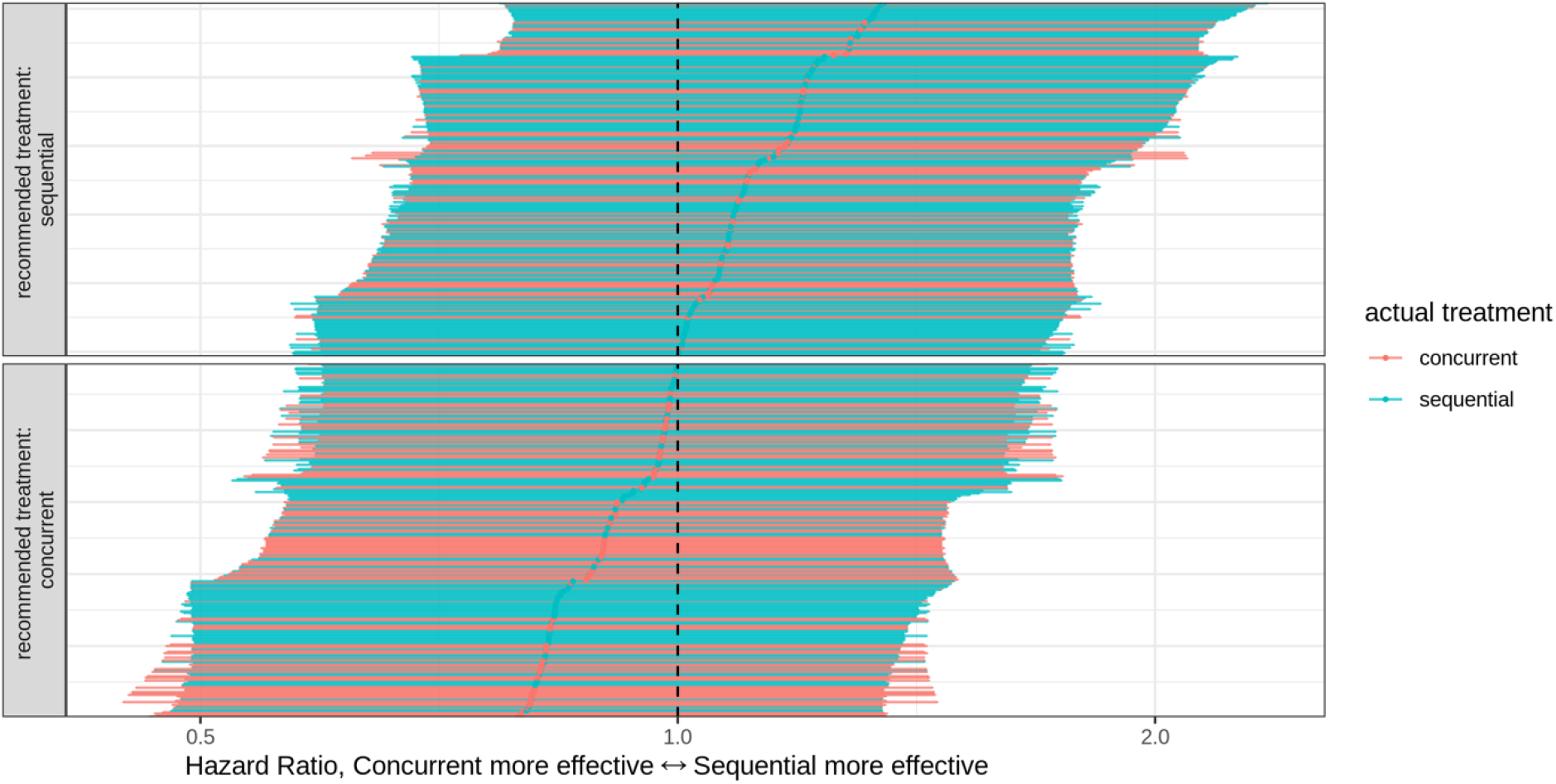
Predicted individual treatment effects for all 504 included patients. Each patient is represented by a horizontal line indicating the 95% credible interval of the predicted hazard ratio for overall survival of concurrent chemoradiation versus sequential chemoradiation, and the point estimate. Colors are added to indicate the actually received treatment. The reference line indicates the null effect: both treatments are equally effective.

As an additional model check, we investigated whether patients with lower estimated overall fitness were more likely to discontinue treatment due to side-effects or disease progression. After estimating patient fitness based on pre-treatment variables, we observed that in patients who underwent concurrent chemoradiation, a lower estimated fitness was associated with a higher probability of discontinuing the original treatment plan after the initiation of treatment (Area Under the Curve of the Receiver Operating Characteristic (AUC), 0.61; 95% confidence interval, 0.55 to 0.68).

### Sampling diagnostics and model fit

We assessed the convergence of the MCMC chains and inspected the possible existence of multiple posterior modes that would prohibit the identification of the joint distribution and thus the treatment effect. The maximum Gelman-Rubin r-hat statistic across all parameters was 1.002 and the posterior density plots were unimodal. Given the large number of independent chains (16) and the number of samples per chain (7500 samples following 2500 warm-up samples) it is unlikely that other posterior modes exist. As outlined earlier, a unimodal posterior distribution implies identification of the treatment effect given the observed data and the modeling assumptions, despite the unobserved confounder. There were 231 divergent transitions in 120,000 samples (0.2%), after inspection of parameter trace plots these were deemed false positives.

The concordance index for overall survival predictions based on pre-treatment variables and the given treatment was 0.59 (95% confidence interval, 0.56 to 0.62). The AUC for predicting the treatment decision based on pre-treatment variables was 0.76 (95% confidence interval, 0.72-0.80).

### Sensitivity analyses

We performed three sensitivity analyses of the average treatment effect estimate. The appendix (section sensitivity analyses) contains additional details on the justification, methods and results of these analyses.

First, we assessed the sensitivity of the average treatment effect estimate to an unmodeled confounder by re-estimating the model with an additional unobserved confounder with a known relationship with treatment and survival. We used several combinations of values for the association between this extra unobserved confounder and treatment and survival. We found that this confounder had to have a confounding strength that was more than half as strong as the modeled unobserved fitness, but opposite in sign with respect to survival, to drive the point estimate of the average treatment effect to a value more extreme than the treatment effect estimate from the RCTs.

Secondly, we assessed the potential bias induced by non-random missing values in weight loss. It is possible that weight loss is more often recorded in the EHR when pronounced weight loss is present. For this specific missingness pattern, treatment effect estimation using the complete cases is unbiased, while imputation may lead to bias *(24)*. This is why we excluded patients with a missing value of weight loss. As weight loss may be related to treatment efficacy and the true prevalence of weight loss is unknown due to the missing data, we recalculated the average treatment effect for several hypothetical values of the prevalence of weight loss by reweighting patients according to their weight loss. In the most extreme hypothetical case where weight loss was always observed if it was present, the average treatment effect estimate (hazard ratio, 1.01; 95% credible interval, 0.65 to 1.53) was very close to the other extreme case where weight loss was missing completely at random (hazard ratio, 1.03; 95% credible interval, 0.71 to 1.68).

Finally, we calculated what the estimated average treatment effect would be when restricting the analysis to a subsample of the cohort that is more like the population of the RCTs *(6)*. Under the assumption that the mechanism for selection for concurrent treatment and the mechanism for selection for inclusion in the RCT are similar, the population was restricted to those with a predicted probability of concurrent treatment higher than several different cut-offs (0%, 25%, 50%, 75%). When restricting the analysis to patients with a higher predicted probability of concurrent treatment to approximate the RCT population, the estimated treatment effect shifted towards concurrent chemoradiation being more effective (hazard ratio 1.01 for predicted probability >0%, N=504; hazard ratio, 1.00 for >25%, N=359; hazard ratio 0.98 for >50%, N=193; hazard ratio 0.95 for >75%, N=55). The treatment effect moves in the direction of the RCT estimate but the estimated survival benefit of concurrent treatment is still smaller. It may be that the method used to match our population with the RCT population was too crude.

## Discussion

We present PROTECT, a method that uses proxy measurements of unobserved confounders to estimate treatment effects from observational data. PROTECT addresses a pervasive problem in observational cancer research: the lack of a direct measurement of the confounder overall fitness. When applied to stage III NSCLC, the results indicate that on average the reported benefit of concurrent over sequential chemoradiation for overall survival may be absent in a real-world population with more patients in lower overall fitness. In our cohort just over half of the patients were treated with sequential chemoradiation, which would imply that in approximately half of the patients the treating physician was confident that concurrent treatment would be beneficial. This statistic fits in with the absence of an average treatment effect. Whereas conventional confounding adjustment methods find a more extreme treatment effect than reported in RCTs, the results from PROTECT are in line with the recommendations from guidelines that patients with lower overall fitness are less likely to benefit from concurrent treatment. This positive association between overall fitness and treatment effect could be due to a higher risk of treatment discontinuation among patients with lower overall fitness when they are assigned the concurrent chemoradiation treatment regimen. Even though the model was not directly optimized to predict discontinuation of treatment, patients for whom the model estimated lower fitness where indeed more likely to discontinue treatment if they were assigned to concurrent treatment. The meta-analysis by Aupérin et al. did not find a statistically significant treatment effect modification by age or ECOG performance score *(6)*. This could be due to the patient inclusion mechanism. All included patients were deemed fit enough for concurrent treatment. This means that older patients included in the RCTs are likely to have been relatively fit for their age. The same principle holds for performance score. If the treatment effect is indeed modified by overall fitness as our results and treatment guidelines suggest, it could be that the variation in fitness is too low in the RCT population to detect the treatment effect modification.

As this is a non-randomized study it is impossible to rule out confounding bias. Several steps were taken to mitigate potential confounding bias. First, we identified potential confounders from literature and domain expertise. We then applied a data-driven model selection procedure that rejects models that do not conform with the confounding structure implicated by the DAG. Lastly, a sensitivity analysis with an independent omitted confounder showed that the results are robust to unobserved confounders of reasonable strength.

Due to the moderate study sample size, our study does not attain high precision in treatment effect estimation. To address this, future studies should be based on data from larger consortia. Furthermore, the discriminatory power of our model for overall survival was low. This could be due to the omission of other important prognostic biomarkers in the analysis, or due to the intrinsic randomness in overall survival time for cancer patients. Our concordance index is in line with a recent meta-analysis of prognostic models for NSCLC patients treated with curative radiotherapy *(25)*.

Most of our patients were treated before approval of durvalumab for stage III NSCLC in the Netherlands. As treatment with durvalumab is contingent on successfully completing the chemotherapy and radiotherapy, the presented model is still of use as the predictions are correlated with successful completion of the treatment regimen. Since durvalumab improves overall survival *(26)*, successfully completing the treatment may become relatively more important than whether the initial treatment was concurrent or sequential chemoradiation.

RCTs remain crucial for treatment effect estimation for cancer patients as they do not suffer from confounding bias. Still there are several situations where treatment effect estimates from observational data are desirable. When parts of the real-world population are not covered by the RCTs for a certain treatment but observational data is available, PROTECT can be used to estimate the treatment effect in these subpopulations. Furthermore, as the method can estimate both average treatment effects and individual treatment effects, PROTECT can be used for studies on biomarkers of treatment efficacy. When new biomarkers become available that were not measured in RCTs, the treatment effect modification of this biomarker can be studied in an observational cohort using PROTECT. In both applications, the resulting estimates may indicate that a new RCT is warranted in specific subpopulations. In this way, observational studies may supplement evidence from RCTs. Conversely, RCTs provide a point of reference for observational studies.

To facilitate future applications of PROTECT, a three-step overview of PROTECT is presented in Table 2. In the appendix (section discussion) we present two examples where PROTECT could be applied, one in unresectable laryngeal carcinoma and one in stage III squamous cell esophageal cancer. In each application there may be additional confounders to consider. However, the core of the PROTECT DAG will be applicable to many different cancer types.

**Table 2.**
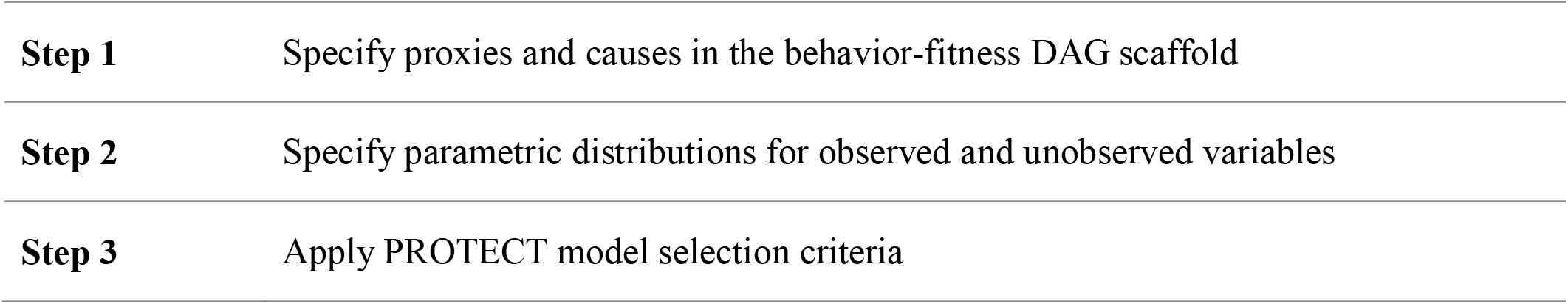
Overview of the PROTECT method for developing individual treatment effect models from observational cancer cohorts. Notes per step: 1. The causal Directed Acyclic Graph (DAG) describes the causal relationships between variables involved in the treatment decisions and outcomes. See Figure 1 for the behavior-fitness DAG scaffold for observational cancer research. To apply PROTECT, researchers need to specify the definitions of treatment, potential additional proxies and causes of tumor behavior and patient fitness, and possibly the outcome. Potential additional application-specific sources of confounding or selection bias must be added as well. 2. These choices should be based on background knowledge. Specific care should be taken to check whether these distributions are uniquely identified. This depends on both the number of proxies of fitness (more is better) and the flexibility of the statistical models. 3. As multiple choices can be made in step 2, applying the PROTECT model selection criteria will reduce the dependence of treatment effect estimates on parameterization choices. Models that do not conform to the confounding structure in the DAG will be rejected. DAG: causal Directed Acyclic Graph; PROTECT: Proxy based individual treatment effect modeling in cancer

|  |  |
| --- | --- |
| **Step 1** | Specify proxies and causes in the behavior-fitness DAG scaffold |
| **Step 2** | Specify parametric distributions for observed and unobserved variables |
| **Step 3** | Apply PROTECT model selection criteria |

In conclusion we present PROTECT, a method for individual treatment effect estimation for cancer patients in the presence of unobserved confounders using proxy measurements. In line with treatment guidelines, our results indicate that sequential treatment should be preferred over concurrent treatment for stage III NSCLC patients who have lower overall fitness. Though on average the two treatments are equivalent in our population, the individual treatment effect estimates of our model can be used to tailor treatment decisions to individual patients.

## Materials and Methods

### Study design

#### Data Source

We conducted a retrospective observational cohort study at the Department of Radiotherapy of the University Medical Center Utrecht, the Netherlands. Patients were referred to the Utrecht center for radiotherapy from the thoracic oncology departments of 9 different hospitals in the Utrecht region in the Netherlands. This study was conducted in accordance with the applicable privacy regulations. As this was a non-experimental retrospective study and most of the patients had died, a waiver for informed consent was obtained from the institutional review board at the University Medical Center Utrecht, along with approval of the study protocol (protocol number WAG/dgv/18/005984). All the methods were performed in accordance with the Declaration of Helsinki.

#### Cohort Selection

Patients referred to our center for the consideration of curative (chemo) radiotherapy as a primary therapy for a first episode of clinical stage III NSCLC between November 2009 and December 2018 were retrospectively identified. Patients had been staged according to the American Joint Committee on Cancer Tumor-Node-Metastasis (TNM) staging protocol. We maintained the TNM version that was clinically used at the time of treatment, spanning versions six *(27)*, seven *(28)* and eight *(29)*. We excluded patients who were eligible for primary surgery or who were treated with palliative intent. Other exclusion criteria were a concurrent other tumor (including a second NSCLC), a prior diagnosis of stage III NSCLC, receiving radiotherapy before chemotherapy to prevent spinal canal invasion, having a Pancoast tumor, receiving radiotherapy at another institution or emigration during follow-up. Patients who were seen at the radiotherapy outpatient clinic but for some reason did not receive radiotherapy were not excluded from the analysis as they are part of the target population for the individual treatment effect model.

#### Definition of intervention and outcome

Clinical variables were extracted from the electronic health records (EHR). Concurrent chemoradiation was defined as a combination of chemotherapy and radiotherapy with time overlap between the treatments, whereas for sequential chemoradiation the start of radiotherapy was subsequent to administration of the last chemotherapy cycle. For both treatments, the planned radiotherapy had to consist of a definitive physical dose of 54 Gray or higher *(30)*. Chemotherapy consisted of two to four cycles of platinum-based chemotherapy (cisplatin or carboplatin, with etoposide, gemcitabine or pemetrexed). As this is a multi-institutional non-experimental study, chemotherapy regimens varied. The goal of an individual treatment effect model is to influence future treatment decisions. Therefore, the intervention under study was concurrent versus sequential chemoradiation according to the initial treatment decision. This choice is in line with the general preference for intention-to-treat analyses in RCTs *(31)*.

The start of follow-up was defined as the date of the last multi-disciplinary tumor board meeting preceding the start of treatment, as this is generally the moment the treatment decision is made.Specific care was taken to record the values of variables as they were known at this time point. The outcome was overall survival measured on a continuous time scale. If not noted in the EHR, data for overall survival was supplemented by querying the Dutch Personal Records Database.

### Statistical Analysis

#### Covariates

The set of variables for the analysis consisted of age, histology (grouped as adenocarcinoma, squamous cell carcinoma, or other), the presence of any weight loss (defined as > 3% of original weight in the 6 months leading up to the treatment decision), performance status 0 versus 1 or higher, defined by the ECOG standard *(32)*, eGFR higher or lower than 60 ml / minute / 1.73 m^2^ and TNM stage IIIA vs IIIB or IIIC.

#### Missing Data Handling

Variables with less than 5% missing values were imputed using mean imputation for continuous variables or a fixed value of 0.5 for binary variables. When the date of the tumor board was unknown this date was imputed based on the date of treatment start with mean imputation per treatment category. Missing values in proxy variables of fitness are assumed to be missing at random conditionally on the observed variables. Further, we assumed that the missingness of weight loss was dependent on the presence of weight loss. For this specific missingness pattern, complete case analysis is unbiased, while imputation may lead to bias *(24)*. Therefore, we excluded patients with a missing value of weight loss. In the appendix (section methods, missing data) we elaborate on this assumption further and present a sensitivity analysis regarding the missingness in weight loss (section sensitivity analyses).

#### Model Evaluation

The treatment effect estimates from PROTECT were contrasted with the baseline approach of including the observed variables in a multivariable Cox-proportional hazards model and an inverse propensity score weighted Cox-proportional hazards model. Estimated treatment effects were compared with the reference value from the meta-analysis by Aupérin *(6)*. As described in the appendix (section methods, pre-processing, parametric models and priors) the model includes a non-linear component. Therefore, potential treatment effect modification for a variable was inspected using partial dependence functions *(23)*.

Model fit for overall survival was assessed using Harrell’s concordance index *(33)*. Model fit for the treatment choice was assessed with the AUC.

Posterior samples were simulated using 16 independently initialized MCMC chains with 7500 samples each, following 2500 warm-up samples. The mixing of chains was inspected with the Gelman-Rubin r-hat statistic *(34)* and the presence of multiple posterior modes was checked visually from posterior density plots.

As an additional model evaluation, we estimated the association between the estimated fitness based on pre-treatment variables and the occurrence of a negative treatment switch anywhere during the treatment. A negative treatment switch was defined as any reduction in treatment intensity compared to the original treatment intention, occurring after the first day of treatment.This included a reduction in chemotherapy dose, fewer chemotherapy cycles, a switch from concurrent to sequential chemoradiation, a lower radiotherapy dose or complete cessation of treatment.

#### Sensitivity analyses

We tested the robustness of the average treatment effect estimate to a hypothetical omitted confounder. This was done by re-estimating the model with an additional unobserved variable with several hypothetical relationships with the treatment and the outcome. Finally, we calculated what the estimated average treatment effect would be when restricting the analysis to a subsample of the cohort that is more like the population of the RCTs *(6)*. Under the assumption that the mechanism for selection for concurrent treatment and the mechanism for selection for inclusion in the RCT are similar, the population was restricted to those with a predicted probability of concurrent treatment higher than several different cut-offs (0%, 25%, 50%, 75%). Details on the justification and implementation of these sensitivity analyses are presented in the appendix (section sensitivity analyses).

#### Implementation

NumPyro version 0.4.1 and JAX version 0.2.7 were used for model estimation. R version 4.0.3 was used for model evaluations.

#### Reporting

For reporting, we adhered to the STROBE reporting guidelines for observational research *(35)*. A completed form is available in the supplemental material.

## Data Availability

Due to local privacy regulations, the original patient data cannot be shared. The code that implements the statistical models and model selection procedure will be made publicly available at an online repository.

RCT: randomized controlled trial
PROTECT: Proxy based individual treatment effect modeling in cancer
NSCLC: non-small cell lung cancer

## Acknowledgements

We kindly acknowledge Dr. Karijn Suijkerbuijk for contributing to the development of the DAG.

## Role of the funding source

WA was supported by the Alexandre Suerman personal PhD stipendium. The Alexandre Suerman stipend had no role in any part of the study design, conduct or reporting.

## Author contributions

Study conception and design: WA, JV, PJ, TL, RE, RR

Corresponding author: WA

Draft manuscript: WA

Data collection and verification: WA, JV, NH, GB

Methodological development: WA, AP, RE, RR

Statistical Analysis: WA, RE, RR

Critical revision and approval of manuscript: all authors

## Competing interests

All authors have completed the ICMJE uniform disclosure form at www.icmje.org/coi_disclosure.pdf and declare: no support from any organization for the submitted work; no financial relationships with any organizations that might have an interest in the submitted work in the previous three years; no other relationships or activities that could appear to have influenced the submitted work; PJ received consulting fees from Sanfit and Inozyme. TL is co-founder and shareholder of Quantib-U B.V.. The department of radiology at the University Medical Center Utrecht has a research collaboration with Philips Healthcare.

## Notes

### Author Declarations

Institutional Review Board of University Medical Center Utrecht waived ethical approval for this work

## References

1. C. M. Booth, I. F. Tannock, Randomised controlled trials and population-based observational research: partners in the evolution of medical evidence, British Journal of Cancer 110, 551–555 (2014).

2. J. H. Lewis, M. L. Kilgore, D. P. Goldman, E. L. Trimble, R. Kaplan, M. J. Montello, M. G. Housman, J. J. Escarce, Participation of Patients 65 Years of Age or Older in Cancer Clinical Trials, JCO 21, 1383–1389 (2003).

3. S. K. Vinod, Decision making in lung cancer – how applicable are the guidelinesã, Clin Oncol (R Coll Radiol) 27, 125–131 (2015).

4. FDA-NIH Biomarker Working Group, Predictive Biomarker (Food and Drug Administration (US), 2016; https://www.ncbi.nlm.nih.gov/books/NBK402283/).

5. D. S. Ettinger, NCCN Non-Small Cell Lung Cancer Guideline, Version 1.2021 (2020) (available at https://www.nccn.org/professionals/physician_gls/pdf/nscl.pdf).

6. A. Aupérin, C. L. Péchoux, E. Rolland, W. J. Curran, K. Furuse, P. Fournel, J. Belderbos, G. Clamon, H. C. Ulutin, R. Paulus, T. Yamanaka, M.-C. Bozonnat, A. Uitterhoeve, X. Wang, L. Stewart, R. Arriagada, S. Burdett, J.-P. Pignon, Meta-analysis of concomitant versus sequential radiochemotherapy in locally advanced non-small-cell lung cancer, J. Clin. Oncol. 28, 2181–2190 (2010).

7. N. Ramnath, T. J. Dilling, L. J. Harris, A. W. Kim, G. C. Michaud, A. A. Balekian, R. Diekemper, F. C. Detterbeck, D. A. Arenberg, Treatment of Stage III Non-small Cell Lung Cancer, Chest 143, e314S–e340S (2013).

8. M. Jamal-Hanjani, G. A. Wilson, N. McGranahan, N. J. Birkbak, T. B. K. Watkins, S. Veeriah, S. Shafi, D. H. Johnson, R. Mitter, R. Rosenthal, M. Salm, S. Horswell, M. Escudero, N. Matthews, A. Rowan, T. Chambers, D. A. Moore, S. Turajlic, H. Xu, S.-M. Lee, M. D. Forster, T. Ahmad, C. T. Hiley, C. Abbosh, M. Falzon, E. Borg, T. Marafioti, D. Lawrence, M. Hayward, S. Kolvekar, N. Panagiotopoulos, S. M. Janes, R. Thakrar, A. Ahmed, F. Blackhall, Y. Summers, R. Shah, L. Joseph, A. M. Quinn, P. A. Crosbie, B. Naidu, G. Middleton, G. Langman, S. Trotter, M. Nicolson, H. Remmen, K. Kerr, M. Chetty, L. Gomersall, D. A. Fennell, A. Nakas, S. Rathinam, G. Anand, S. Khan, P. Russell, V. Ezhil, B. Ismail, M. Irvin-Sellers, V. Prakash, J. F. Lester, M. Kornaszewska, R. Attanoos, H. Adams, H. Davies, S. Dentro, P. Taniere, B. O’Sullivan, H. L. Lowe, J. A. Hartley, N. Iles, H. Bell, Y. Ngai, J. A. Shaw, J. Herrero, Z. Szallasi, R. F. Schwarz, A. Stewart, S. A. Quezada, J. Le Quesne, P. Van Loo, C. Dive, A. Hackshaw, C. Swanton, Tracking the Evolution of Non–Small-Cell Lung Cancer, New England Journal of Medicine 376, 2109–2121 (2017).

9. J. Pearl, Ed., in Causality, (Cambridge University Press, Cambridge, 2009), pp. 65–106.

10. S. Greenland, The Effect of Misclassification in the Presence of Covariates, American Journal of Epidemiology 112, 564–569 (1980).

11. E. Ogburn, T. Vanderweele, Bias attenuation results for nondifferentially mismeasured ordinal and coarsened confounders, Biometrika 100, 241–248 (2013).

12. M. Kuroki, J. Pearl, Measurement bias and effect restoration in causal inference, Biometrika 101, 423–437 (2014).

13. P. R. Rosenbaum, D. B. Rubin, The central role of the propensity score in observational studies for causal effects, Biometrika 70, 41–55 (1983).

14. W. Miao, Z. Geng, E. T. Tchetgen, Identifying Causal Effects With Proxy Variables of an Unmeasured Confounder, (2016).

15. N. Kallus, X. Mao, M. Uehara, Causal Inference Under Unmeasured Confounding With Negative Controls: A Minimax Learning Approach, 2103.14029 [cs, stat] (2021) (available at http://arxiv.org/abs/2103.14029).

16. Y. Wang, D. M. Blei, The Blessings of Multiple Causes, Journal of the American Statistical Association 114, 1574–1596 (2019).

17. S. Lee, E. Bareinboim, Causal Identification with Matrix Equations, Columbia CausalAI Laboratory Technical Report (R-70) (2021).

18. C. Louizos, U. Shalit, J. Mooij, D. Sontag, R. Zemel, M. Welling, Causal Effect Inference with Deep Latent-Variable Models, (2017).

19. M. D. Hoffman, A. Gelman, The No-U-Turn Sampler: Adaptively Setting Path Lengths in Hamiltonian Monte Carlo, 1111.4246 [cs, stat] (2011) (available at http://arxiv.org/abs/1111.4246).

20. D. Phan, N. Pradhan, M. Jankowiak, Composable Effects for Flexible and Accelerated Probabilistic Programming in NumPyro, arXiv preprint 1912.11554 (2019).

21. J. Bradbury, R. Frostig, P. Hawkins, M. J. Johnson, C. Leary, D. Maclaurin, G. Necula, A. Paszke, J. VanderPlas, S. Wanderman-Milne, Q. Zhang, JAX: composable transformations of Python+NumPy programs (2018; http://github.com/google/jax).

22. K. Burke, M. C. Jones, A. Noufaily, A Flexible Parametric Modelling Framework for Survival Analysis, Journal of the Royal Statistical Society: Series C (Applied Statistics) 69, 429–457 (2020).

23. J. H. Friedman, Greedy function approximation: A gradient boosting machine., Ann. Statist. 29, 1189–1232 (2001).

24. I. R. White, J. B. Carlin, Bias and efficiency of multiple imputation compared with complete-case analysis for missing covariate values, Statistics in Medicine 29, 2920–2931 (2010).

25. G. Kothari, J. Korte, E. J. Lehrer, N. G. Zaorsky, S. Lazarakis, T. Kron, N. Hardcastle, S. Siva, A systematic review and meta-analysis of the prognostic value of radiomics based models in non-small cell lung cancer treated with curative radiotherapy, Radiotherapy and Oncology 155, 188–203 (2021).

26. C. Faivre-Finn, D. Vicente, T. Kurata, D. Planchard, L. Paz-Ares, J. F. Vansteenkiste, D. R. Spigel, M. C. Garassino, M. Reck, S. Senan, J. Naidoo, A. Rimner, Y.-L. Wu, J. E. Gray, M. Özgüroğlu, K. H. Lee, B. C. Cho, T. Kato, M. de Wit, M. Newton, L. Wang, P. Thiyagarajah, S. J. Antonia, Four-Year Survival With Durvalumab After Chemoradiotherapy in Stage III NSCLC—an Update From the PACIFIC Trial, Journal of Thoracic Oncology 16, 860–867 (2021).

27. TNM Atlas, 6th Edition | WileyWiley.com (available at https://www.wiley.com/en-nl/TNM+Atlas%2C+6th+Edition-p-9781118695609).

28. TNM Classification of Malignant Tumours, 7th Edition | WileyWiley.com (available at https://www.wiley.com/en-nl/TNM+Classification+of+Malignant+Tumours%2C+7th+Edition-p-9781444358964).

29. TNM Classification of Malignant Tumours, 8th Edition | WileyWiley.com (available at https://www.wiley.com/en-us/TNM+Classification+of+Malignant+Tumours%2C+8th+Edition-p-9781119263579).

30. S. J. Antonia, A. Villegas, D. Daniel, D. Vicente, S. Murakami, R. Hui, T. Yokoi, A. Chiappori, K. H. Lee, M. de Wit, B. C. Cho, M. Bourhaba, X. Quantin, T. Tokito, T. Mekhail, D. Planchard, Y.-C. Kim, C. S. Karapetis, S. Hiret, G. Ostoros, K. Kubota, J. E. Gray, L. Paz-Ares, J. de Castro Carpeño, C. Wadsworth, G. Melillo, H. Jiang, Y. Huang, P. A. Dennis, M. Özgüroğlu, PACIFIC Investigators, Durvalumab after Chemoradiotherapy in Stage III Non-Small-Cell Lung Cancer, N. Engl. J. Med. 377, 1919–1929 (2017).

31. S. K. Gupta, Intention-to-treat concept: A review, Perspect Clin Res 2, 109–112 (2011).

32. M. M. Oken, R. H. Creech, D. C. Tormey, J. Horton, T. E. Davis, E. T. McFadden, P. P. Carbone, Toxicity and response criteria of the Eastern Cooperative Oncology Group, American Journal of Clinical Oncology 5, 649–656 (1982).

33. F. E. Harrell, R. M. Califf, D. B. Pryor, K. L. Lee, R. A. Rosati, Evaluating the Yield of Medical Tests, JAMA 247, 2543–2546 (1982).

34. A. Vehtari, A. Gelman, D. Simpson, B. Carpenter, P.-C. Bürkner, Rank-normalization, folding, and localization: An improved $\widehat{R}$ for assessing convergence of MCMC, Bayesian Anal. (2020), doi:10.1214/20-BA1221.

35. E. von Elm, D. G. Altman, M. Egger, S. J. Pocock, P. C. Gøtzsche, J. P. Vandenbroucke, STROBE Initiative, The Strengthening the Reporting of Observational Studies in Epidemiology (STROBE) statement: guidelines for reporting observational studies, Ann Intern Med 147, 573–577 (2007).

