## appendix for "Individual treatment effect estimation in the presence of unobserved confounding using proxies: a cohort study in stage III non-small cell lung cancer"

Wouter A.C. van Amsterdam et al

### Appendix

#### A Methods

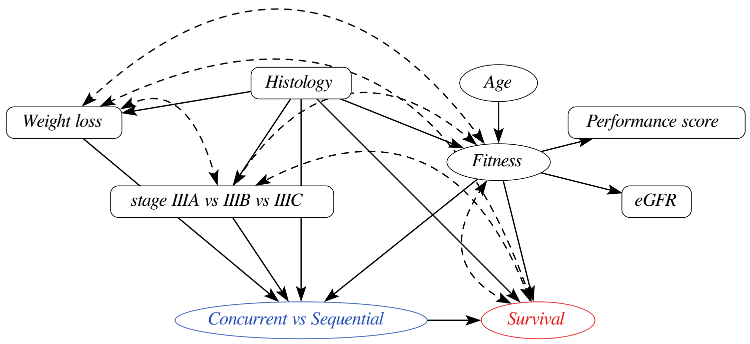

Figure 1: Acyclic Directed Mixed Graph, resulting from marginalizing out $TumorBehavior$ in the original DAG. A dashed bi-directed arrow indicates the presence of an unobserved common cause ($TumorBehavior$). eGFR: estimated Glomerular Filtration Rate

#### A.1 Treatment Effect Estimation with PROTECT

In this section we introduce additional implementation details of the PROTECT method.

##### A.1.1 Marginalizing out Tumor Behavior for Estimation

The PROTECT DAG has two latent factors: tumor behavior and patient fitness. Since the latent factors are not observed they need to be marginalized out, both during the model estimation phase and for posterior predictions on new data. As conditioning on the latent factor for tumor behavior is not needed to satisfy the backdoor rule, we can also do inference over the acyclic directed mixed graph (ADMG) [18] that results from marginalizing out tumor behavior, without harming the ability to estimate the conditional treatment effect. ADMGs are a generalization of DAGs that allow for bi-directed arrows, indicating the presence of an unobserved confounder [18]. The AMDG that results from marginalizing out tumor behavior has fewer parameters to estimate and was therefore used for model estimation. See Figure A.1.1 for the resulting ADMG. There are multiple bi-directed arrows in the ADMG as a result of marginalizing out tumor behavior. To model these dependencies, the original ancestral order is used. This means for instance that the outcome will be modeled conditional on stage and not vice-versa, as stage is an ancestor of the outcome in both the original DAG and the ADMG with tumor behavior marginalized out.

##### A.1.2 Target Estimand

Here we derive a non-parametric expression for estimating the model from the observed data. Let $t_x \in \{0, 1\}$ be the treatment indicator where. Let $y$ denote the outcome. Let $X$ and $W$ be vectors of covariates (causes and proxies of fitness respectively, in line with the AMDG in Figure A.1.1). Let $X^B$ denote the proxies and causes of tumor behavior from the original ADMG (histology, weight loss and stage IIIA vs IIIB vs IIIC in the case of stage III Non-Small Cell Lung Cancer). We use Pearl's do-operator to indicate intervening on a variable [16]. We are interested in the conditional average treatment effect (CATE) as the difference in expected survival under the different treatments:

$$\text{CATE}(X, W, X^B) = \mathbf{E}\left[y|\text{do}(t_x = 1), X, W, X^B\right] - \mathbf{E}\left[y|\text{do}(t_x = 0), X, W, X^B\right] \tag{1}$$

The presented AMDG suggests a causal factorization of the conditional distribution in equation (1). We will now derive our target estimand of the joint distribution of the observable variables $(y, t_x, W)$ given the control variables $(X, X^B)$.

*Proof.* Let $y$ be the outcome, $t_x$ the treatment variable, $F \in \mathbb{R}$ a latent factor, $W$ possibly multidimensional proxy variables for $F$, and $X$ possibly multidimensional causes for $F$, $X^B$ possibly multidimensional causes and proxies for tumor behavior, then

$$p(y, t_x, W|X, X^B)$$

$$=^a \int_F p(y, t_x, W|X, X^B, F)p(F|X, X^B)dF \tag{2}$$

$$=^b \int_F p(y|t_x, W, X, X^B, F)p(t_x|W, X, X^B, F)p(W|X, X^B, F)p(F|X, X^B)dF \tag{3}$$

$$=^c \int_F p(y|t_x, X^B, F)p(t_x|X^B, F)p(W|F)p(F|X, X^B)dF \tag{4}$$

□

a) by the law of total probability

b) by the chain rule of of probability

c) by conditional independencies implied by the ADMG

In 4 the outcome distribution conditions on all confounders, thereby satisfying the backdoor rule. This implies we can use Rule 2 of the rules of do-calculus and exchange observing $t_x$ (as we do in the observational distribution from which our samples are drawn), with intervening on $t_x$ (the interventional distribution that is the target of our research) [15].

After estimating the joint distribution, we can calculate the CATE for a patient by conditioning 4 on the observed proxy variables $W$ for both $t_x = 0$ and $t_x = 1$ and marginalizing over $F$, and then calculating the difference in the expected value of $y$ for $t_x = 1$ and $t_x = 0$. The average treatment effect can be estimated by calculating the mean CATE over all observed patients.

##### A.1.3 Estimation in a Marginalized DAG

For any application of PROTECT, the number of possible cause and proxy variables of patient fitness may be large. Moreover, different research groups investigating the same application may come up with different sets of cause and proxy variables of fitness. A natural question is whether the estimated treatment effect depends on the chosen set of proxies. Under the assumption that there is a finite number of potential cause and proxy variables of fitness, we will show that omitting some these variables will not necessarily bias the treatment effect estimate. For this we need two things to hold:

1. The DAG with fewer variables is indeed the DAG that is obtained after marginalizing out the unused variables from the full DAG
2. Given the DAG with fewer variables, the conditional average treatment effect is identified from the observed data

We defer the discussion of the second requirement to subsection A.1.3. To demonstrate the first requirement we will consider a hypothetical application of PROTECT to a case where there exist three binary proxies of fitness $(w_1, w_2, w_3)$ and no cause variables for fitness. A research groups tries to estimate the treatment effect but only has measurements of two of the three proxy variables. See Figure A.1.3 for the DAG for this situation.

As the researchers only have samples from

$$p(y, t_x, w_1, w_2) = \sum_{\mathrm{w}=0}^{1} p(y, t_x, w_1, w_2, w_3 = \mathrm{w})$$

the target estimand for this study is $\mathrm{CATE}(w_1, w_2)$ instead of $\mathrm{CATE}(w_1, w_2, w_3)$. If the group had access to the joint distribution $p(y, t_x, w_1, w_2, w_3)$, they could calculate $\mathrm{CATE}(w_1, w_2)$ by first summing out $w_3$ from the joint distribution and then calculating the CATE. The question is whether this will lead to the same estimand as when estimating $\mathrm{CATE}(w_1, w_2)$ from samples from $p(y, t_x, w_1, w_2)$ directly. We now show that this is indeed the case.

$$\begin{aligned}
&\sum_{\mathrm{w}=0}^{1} p(y, t_x, w_1, w_2, w_3 = \mathrm{w}) \\
&= \sum_{\mathrm{w}=0}^{1} \int_F p(y|t_x, F) p(t_x|F) p(w_1|F) p(w_2|F) p(w_3 = \mathrm{w}|F) p(F) dF \\
&= \int_F p(y|t_x, F) p(t_x|F) p(w_1|F) p(w_2|F) \sum_{\mathrm{w}=0}^{1} \left[p(w_3 = \mathrm{w}|F)\right] p(F) dF \\
&=^1 \int_F p(y|t_x, F) p(t_x|F) p(w_1|F) p(w_2|F) p(F) dF \\
&= p(y, t_x, w_1, w_2)
\end{aligned}$$

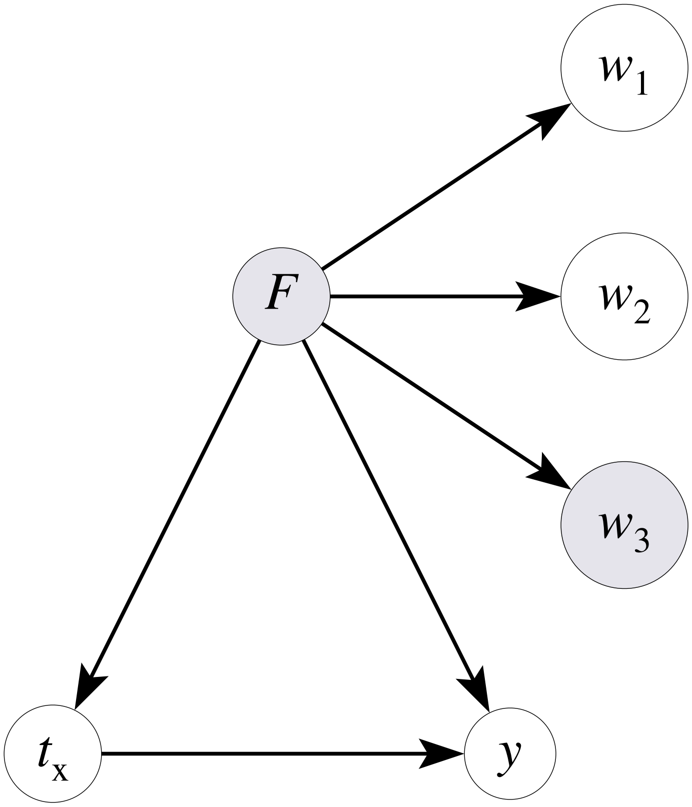

Figure 2: Example DAG where one of the three proxies is not observed.

In [1] the back-door requirement is still fulfilled so the treatment effect can be estimated. The average treatment effect is calculated by calculating the expectation of the CATE over the proxy variables. As the CATE$(w_1, w_2)$ can be estimated from samples of $p(y, t_x, w_1, w_2)$ and the order of taking the expectation over proxy variables does not matter, the inferred average treatment effect would be the same as first estimating CATE$(w_1, w_2, w_3)$ and then taking the expectation over all three proxy variables. A similar argument can be made for cause variables of $F$.

###### A.1.4 Treatment Effect Estimation

As mentioned in the main text, if the joint distribution of observed variables and the latent confounder can be estimated from the observed data, the treatment effect can be estimated. This is because the back-door adjustment formula can be applied using the estimated distributions of survival given treatment and fitness, and fitness given the proxy variables and cause variables of fitness [15, 11], see also equation 4. The crucial question is whether the joint distribution of observed variables and the latent confounder can be correctly estimated from the observed data. Without constraints on the joint distribution, this is not possible. Fortunately, the DAG provides conditional independency constraints on the joint distribution. For instance, the treatment choice is independent of performance score, once the value of fitness is known. However, this is not enough to identify the joint distribution in general and additional assumptions are needed. These assumptions can be provided by the form of error distributions of the observed variables and latent confounder or by functional forms of relationships between these variables.

In PROTECT, parametric forms for the structural equations in the DAG are specified. By assuming parametric models and thus reducing the family of structural causal models under consideration, the question of identification of the treatment effect reduces to whether the parameters for the structural equations can be uniquely estimated from observed data. This is not an easy question to answer in general. If all observed variables and the latent factor followed linear models with Gaussian error distributions, a well-known sufficient condition for uniqueness of the parameters is when there are at least three independent proxies per latent factor [2]. This textbook result of parameter identification is based on comparing the number of unknown parameters in the statistical model with the number of unique entries in the covariance matrix of the observed variables. The latent factor fitness in our DAG has four dependent variables (two proxies, the treatment and overall survival). Treatment and survival also have a direct dependency relationship that would require one additional parameter to be estimated in a linear Gaussian setting. The requirement of estimating this single extra parameter is offset by having one more observed variable, so this model would still be identified. However, many settings will be more complex than standard linear structural equation models, and analytical identification proofs are often intractable to obtain [19]. Instead, empirical checks can be performed to see if there is any evidence of non-uniqueness of the treatment

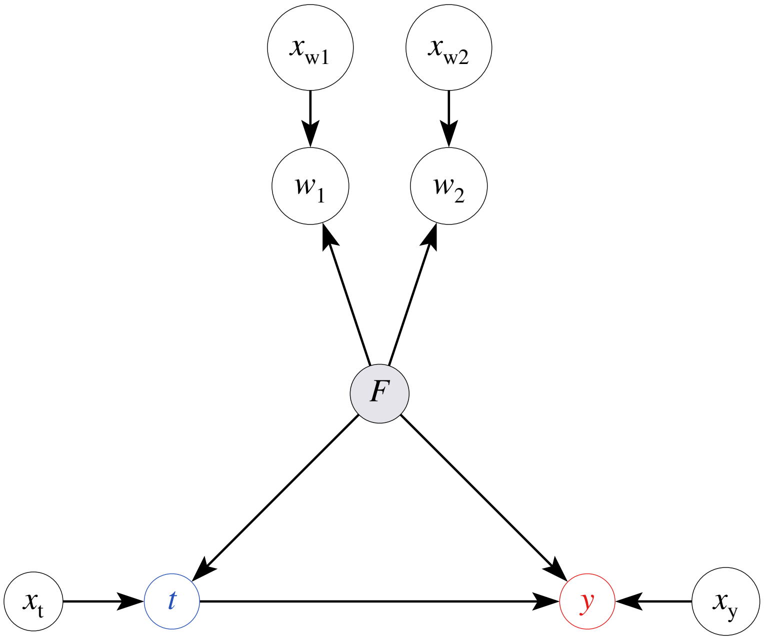

Figure 3: Exeample of a DAG where PROTECT could be applied

effect estimate given the observed data and modeling assumptions. In a Bayesian parameter estimation setting, this amounts to checking the posterior distribution over parameters for multi-modality.

###### A.1.5 Model Selection

In PROTECT, the joint distribution of observed variables and unobserved variables is estimated by specifying parametric distributions for all structural equations implied by the DAG. Different parameterizations may lead to different inferences regarding the treatment effect. To reduce the dependence of the treatment effect estimate on these choices, we now introduce a data-driven model selection procedure. We describe it for a general DAG with a latent confounder with proxy variables and causes.

In figure 3 we present a general latent confounder model with two proxies $w_1, w_2$, a treatment variable $t$ and outcome $y$. Each observed variable has a possibly empty set of control variables $\mathbf{x}_{(.)}$ and the sets of control variables are allowed to overlap. Causes of the latent factor $F$ are permitted but are irrelevant to the model selection procedure so they are omitted here. The latent factor $F$

is a cause of the proxies, treatment and outcome, and is the only confounder of $t$ and $y$. The proxies, treatment and outcome are assumed to be random variables conditional on their parents in the graph. This graph is necessarily an abstraction of complicated clinical and biological processes. In reality, the process that is responsible for treatment decisions and outcomes may be better described with a multi-dimensional latent factor $F$. Considering this multi-dimensional $F$, it is likely that there are dimensions of $F$ that are related to treatment but not to the outcome ($F_t$) and vice-versa ($F_y$). The dimensions of $F$ that are related to both treatment and outcome are denoted $F_{t,y}$ and constitute the true confounder. See Figure 4. Possible clinical interpretations of these dimensions are:

- $F_t$: information that the pre-treatment variables provide that is thought to be relevant to efficacy of treatment (and is used as such to select patients for treatment), but in reality holds no information on treatment efficacy or outcome
- $F_y$: information that the pre-treatment variables have on the outcome that is not known to the physician (this could be treatment effect modification that is not known to the physician), or otherwise is not used to make treatment decisions.
- $F_{t,y}$: information that the pre-treatment variables have on the outcome or treatment efficacy and that is utilized in the treatment decision process

To estimate the treatment effect, the modeled latent factor $\hat{F}$ should contain as much information on $F_{t,y}$ as possible. Both $F_t$ and $F_y$ are irrelevant for the estimation of the treatment effect in terms of bias.

When the parameterization of the model has limited expressiveness (e.g. $\hat{F}$ is a one-dimensional latent variable), it is not guaranteed that the model parameters that maximize the joint likelihood of the observed data will learn an $\hat{F}$ that has information about $F_{t,y}$. For example, when the mutual information between the pre-treatment variables and treatment is much higher than the mutual information between pre-treatment variables and outcome, it could be that a model with a single dimensional $\hat{F}$ will learn $\hat{F} = F_t$. Indeed, this is not an unrealistic scenario. The treatment selection process is mostly determined by the pre-treatment variables. Outcomes like overall survival have much higher intrinsic randomness given these pre-treatment variables as they depend on 1. the true biological state of the patient and the tumor that may never be fully known and 2. random events in the future that have not occurred at the time of the treatment choice. In the following paragraph we introduce a set of model checks based on observed data to evaluate whether a given parameterization of the graphical model in the DAG is compatible with the assumptions laid out in the DAG and the requirement of modeling $\hat{F} = F_{t,y}$

**Hyperparameter Selection** When there are multiple choices for parameterizing the joint model summarized in hyperparameter $\psi$ (e.g. number of

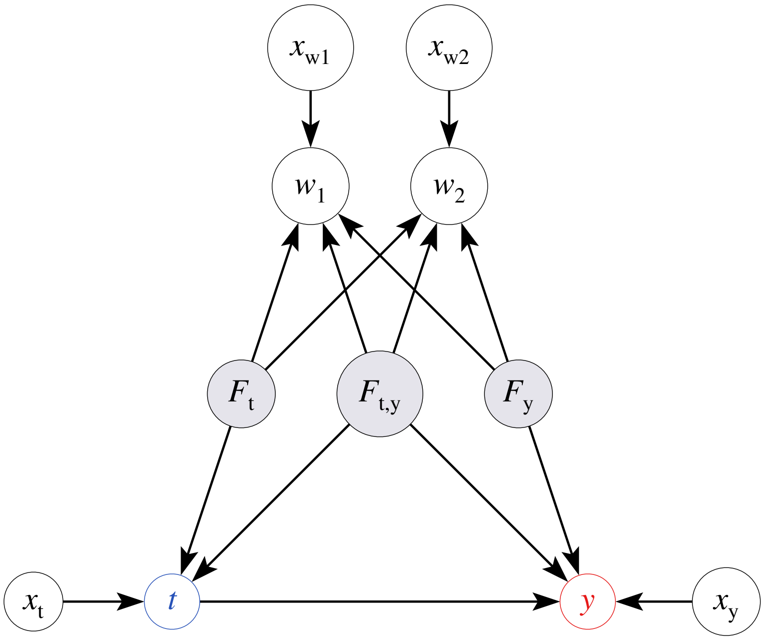

Figure 4: A more complex true data generating process may underlie the observed data which is modeled as in Figure 3 $F_t$ is a latent factor that only influences the proxy variables and the treatment assignment. $F_y$ is a latent factor that only influences the proxy variables and the outcome. $F_{t,y}$ is a latent factor that influences the proxy variables and both the treatment and the outcome. $F_{t,y}$ is the only confounder.

dimensions for $\hat{F}$, parameterization choices for any of the conditional distributions, including priors for parameters), each value of $\psi$ will imply a posterior distribution $p_{\text{post},\psi}(\theta) = p_\psi(\theta|D_{\text{train}})$ over global parameters $\theta$ after conditioning on training data $D_{\text{train}}$. To infer if a chosen hyperparameter $\psi$ is compatible with the assumptions on the data generating mechanism in the DAG, a number necessary constraints implied by the DAG can be checked on held-out data $D_{\text{test}}$. Let $p_\psi(y^{(i)}|t^{(i)}, \mathbf{w}^{(i)}, \mathbf{x}^{(i)}, \theta)$ be the predictive density for the outcome of observation $i$, conditional on treatment, proxies and control variables for the outcome, of the model with hyperparameter $\psi$ and global parameter value $\theta$. Let $l_\psi(y^{(i)}|t^{(i)}, \mathbf{w}^{(i)}, \mathbf{x}^{(i)})$ be the log point-wise predictive density for observation $i$ from the test data, given hyperparameter value $\psi$: $l_\psi(y^{(i)}|t^{(i)}, \mathbf{w}^{(i)}, \mathbf{x}^{(i)}) = \log \mathbb{E}_{\theta \sim p_{\text{post},\psi}(\theta)} p_\psi(y^{(i)}|t^{(i)}, \mathbf{w}^{(i)}, \mathbf{x}^{(i)}, \theta)$. Here the expectation over $\theta$ can be approximated e.g. by using monte-carlo samples of $\theta$ from the posterior distribution. Let $L_\psi(y^{(i)}|t^{(i)}, \mathbf{w}^{(i)}, \mathbf{x}^{(i)})$ be the expectation of the log point-wise predictive likelihood (*elpd*) [21] over the held-out data, and let the *elpd* for different conditional distributions be similarly defined, then for $t, y$ and proxies $w_j$ the following minimal criteria should hold:

$$L_\psi(w_j|y, t, \mathbf{w}_{-j}, \mathbf{x}_{w_j}) = \mathbb{E}_{F \sim p_\psi(F|y,t,\mathbf{w}_{-j},\mathbf{x}_{w_j})} p_\psi(w_j|\mathbf{x}_{w_j}, F) > L(w_j|\mathbf{x}_{w_j}) \quad (5)$$

$$L_\psi(t|y, \mathbf{w}, \mathbf{x}_t) = \mathbb{E}_{F \sim p_\psi(F|y,\mathbf{w},\mathbf{x}_t)} p_\psi(t|\mathbf{x}_t, F) > L(t|\mathbf{x}_t) \quad (6)$$

$$L_\psi(y|t, \mathbf{w}, \mathbf{x}_y) = \mathbb{E}_{F \sim p_\psi(F|t,\mathbf{w},\mathbf{x}_y)} p_\psi(y|t, \mathbf{x}_y, F) > L(y|t, \mathbf{x}_y) \quad (7)$$

In words this means that for each observed variable, the predictive likelihood of the model with the latent variable that conditions on the other observed variables should be greater than a corresponding baseline model that conditions only on the direct parents in the DAG, excluding the latent factor. To calculate the *elpd* for the baseline models, a separate regression model was formulated for each target using the same linear formula, outcome distribution and priors as in the DAG model, but without the latent factor. If the predictive likelihood of the model with latent factor is higher than the baseline regression model without the latent factor, then the latent factor effectively transmits information between observed variables in the model indexed by $\psi$. However, this still does not guarantee that the inferred $\hat{F}$ is the actual confounder $F_{t,y}$. If we select a hyperparameter $\psi = \psi_t$ that leads to $\hat{F} = F_t$ then equations 5 may still be satisfied. We can formulate more strict requirements. If we mutilate the DAG in Figure 4 by removing $F_{t,y}$ and $F_y$ and their outgoing arrows, the following equality can be shown by applying the conditional independence of $F_t|t$ of $y|t$ implied by the mutilated DAG:

$$\begin{aligned} p_{\psi_t}(w_j|y, t, \mathbf{w}_{-j}, \mathbf{x}_{w_j}) &= \mathbb{E}_{F_t \sim p_\psi(F_t|y,t,\mathbf{w}_{-j},\mathbf{x}_{w_j})} p_\psi(w_j|\mathbf{x}_{w_j}, F_t) \\ &= \mathbb{E}_{F_t \sim p_\psi(F_t|t,\mathbf{w}_{-j},\mathbf{x}_{w_j})} p_\psi(w_j|\mathbf{x}_{w_j}, F_t) \\ &= p_{\psi_t}(w_j|t, \mathbf{w}_{-j}, \mathbf{x}_{w_j}) \end{aligned}$$

Equivalently, if we select a hyperparameter $\psi = \psi_y$ that leads to $\hat{F} = F_y$ then $L_{\psi_y}(w_j|y, t, \mathbf{w}_{-j}, \mathbf{x}_{w_j}) = L_{\psi_y}(w_j|y, \mathbf{w}_{-j}, \mathbf{x}_{w_j})$. Note that this equality does not hold if either $F_{t,y}$, or the combination of $F_t$ and $F_y$ are in the DAG. We can now define a necessary condition for the $\hat{F}$, estimated from the observed data, not to be independent of $F_{t,y}$. For all $w_j$:

$$L_{\psi_{t,y}}(w_j|y, t, \mathbf{w}_{-j}, \mathbf{x}_{w_j}) > L_{\psi_{t,y}}(w_j|t, \mathbf{w}_{-j}, \mathbf{x}_{w_j}) \quad (8)$$

$$L_{\psi_{t,y}}(w_j|y, t, \mathbf{w}_{-j}, \mathbf{x}_{w_j}) > L_{\psi_{t,y}}(w_j|y, \mathbf{w}_{-j}, \mathbf{x}_{w_j}) \quad (9)$$

Hyperparameter settings for which one of these conditions does not hold for one of the proxies should be rejected. Note that this does not rule out the possibility that $\hat{F}$ is some function of $F_t$ and $F_y$ and is still not related to $F_{t,y}$.

#### A.2 Methods for stage III NSCLC application

In this subsection more implementation details for the application of PROTECT to stage III Non-Small Cell Lung cancer (NSCLC) are provided.

##### A.2.1 Missing Data

**Proxies of fitness** Values of proxies of fitness are assumed to be missing at random conditional on the observed variables. We further assumed independent priors for the proxy missingness mechanism. Together with the missing at random assumption, this makes the missingness model ignorable, meaning that it does not contribute to the estimation of the other parameters. We therefore did not model missingness in proxies. Since we model the joint distribution of proxies, treatment and survival, marginalization of the missing values of proxy variables is trivial by not adding terms to the likelihood for values that are not observed.

**Weight loss** Roughly 15% of the values for weight loss were missing. Though weight loss is a known prognostic factor and is a standard part of the pre-treatment history taking, the physician may sometimes forget to ask about it, or forget to note it down in the electronic health record. It is likely that high values of weight loss have a higher probability of being registered. A patient with high weight loss may self report it and the physician is more likely to report it, as it is more notable. This renders the missingness mechanism for weight loss "Missing Not at Random". When the missingness in a covariate is dependent on the value of the covariate (but not on the outcome), estimating parameters of a regression model for the outcome using the complete cases is not biased by the missingness, while imputation may lead to bias in the estimation of the treatment effect [22]. A problem with the complete case method here is that allthough the conditional treatment effect may be validly estimated, the average treatment effect may be different in the population of interest than in the complete case population, as the populations may differ with respect to the distribution of weight loss. If the effect of treatment depends on weight loss (according to our ADMG, either through the effect of tumor behavior on survival, or through the effect of tumor behavior on fitness), the average treatment effect estimated from the complete cases with respect to weight loss is a biased estimator for the average treatment effect in the target population. Through a sensitivity analysis presented in section B.2.2 we assess what the effect of this missingness on the estimated average treatment effect may be.

##### A.2.2 Pre-processing, Parametric Models and Priors

In this subsection we describe details on the data pre-processing, the parametric forms of each conditional distribution and the priors for all parameters. The Gaussian distribution parameterized with mean $\mu$ and standard deviation $\sigma$ is denoted $\mathcal{N}(\mu, \sigma)$, the Half-Normal distribution with standard deviation $\sigma$ is denoted HN($\sigma$), the Bernoulli distribution with probability $p$ is denoted as Bern($p$), the Uniform distribution over values between $a$ and $b$ is denoted $\mathcal{U}(a, b)$

**Covariate pre-processing** In order to be able to use the same distribution to model each of the proxies, proxy variables were binarized. Dichotomization of variables is not generally recommended due to the loss of information. However, as our primary goal is to identify the conditional average treatment effect, we expect the potential loss of information from the dichotomization to be small compared to the extra modeling challenges when dealing with proxies with different outcome distributions. Our exact variable definitions were:

- ECOG performance score: 1 or higher vs 0 (higher means worse overall health according to the ECOG standard [14])
- eGFR: less (or greater) than 60 milliliter / minute / $1.73m^2$

The first value (1 for ECOG, and less than 60 for eGFR) was dummy coded as 0, and the second value was dummy coded as 1. There were only two patients with stage IIIC, therefore they were grouped with IIIB patients. Histology types were grouped as Adenocarcinoma, Squamous Cell Carcinoma or other, missing values were classified as other. Weight loss was defined as any weight loss of $\geq 3\%$ of the original weight in the 6 months preceding the start of follow-up. Age was scaled to zero mean and standard deviation 1. Details on the definition of the treatment variable and outcome are given in the main text.

**Survival** The Cox-proportional hazards model is characterized by a partial likelihood that leaves the baseline hazard unspecified. Performing Bayesian inference with a proportional hazards model will require specifying a likelihood for the baseline hazard. We chose a parametric survival model, using the Adaptive Power Generalized Weibull (APGW) distribution as described in [4]. The APGW has two parameters that control the shape of the baseline hazard function. With these two parameters, the APGW can model a wide range of baseline hazard function shapes. The APGW accommodates non-proportional hazards effects by letting one or more of the shape parameters depend on covariates. It can be parameterized as an accelerated failure-time model or as a (proportional) hazards model. Using the proportional hazards formulation of the APGW makes it very similar to Cox-proportional hazards regression, but with a parametric baseline hazard function. The $\beta$ coefficients in this version have the same interpretation as the parameters in the familiar Cox-proportional hazards regression, namely log hazard ratios. The reference value for the treatment effect is presented as a hazard ratio [1]. This value assumes a proportional hazard model for overall survival. Therefore, we chose to parameterize the outcome model similarly using a linear model for the log-hazard ratio in the proportional hazards formulation of the APGW. Potential treatment effect modification for variables was accommodated by adding parameters for product terms of the variable and the treatment. The APGW was parameterized as follows:

$$\beta(t_x, F, X_y) = \beta_0 + F\beta_{F\to y} + X_y\beta_{X_y\to y} + \tag{10}$$
$$t_x(\beta_{t_x\to y} + F\beta_{F*t_x\to y} + X_y\beta_{X_y*t_x\to y}) \tag{11}$$
$$\{\text{time}, \text{deceased}\} \sim \text{APGW}(0, \beta(t_x, F, X_y), \alpha_0, \nu_0) \tag{12}$$
$$\beta_{X_y\to y}, \beta_{t_x\to y} \sim \mathcal{N}(0, 2.5) \tag{13}$$
$$\beta_0, \alpha_0 \sim \mathcal{N}(0, 5) \tag{14}$$
$$\nu_0 \sim \mathcal{U}(-5, 5) \tag{15}$$
$$\beta_{F\to y} \sim \mathcal{N}(0, \sigma_{F\to y}) \tag{16}$$
$$\beta_{X_y*t_x\to y}, \beta_{F*t_x\to y} \sim \mathcal{N}(0, 0.1) \tag{17}$$

Here, $t_x = 1$ is concurrent chemoradiation, $F$ is the inferred latent fitness, $X_y$ are the other direct parents of survival in the ADMG, $\sigma_{F\to y}$ is a hyperparameter that was determined per the model selection method in subsection A.1.5. Using this parameterization, we can now express the CATE (equation 1) for patient $i$ in terms of a log hazard ratio $\beta$:

$$\text{CATE}^{(i)} = \beta(t_x = 1, F^{(i)}, X_y^{(i)}) - \beta(t_x = 0, F^{(i)}, X_y^{(i)}) \tag{18}$$
$$= \beta_{t_x\to y} + F^{(i)}\beta_{F*t_x\to y} + X_y^{(i)}\beta_{X_y*t_x\to y} \tag{19}$$

The average treatment effect (ATE) is estimated with the mean CATE over the observed population.

**Latent Factor** The latent factor $F$ is modeled using a conditional Gaussian distribution with fixed standard deviation 1, followed by the logistic function (also known as the sigmoid function, denoted $\sigma$). The logistic function was used to fix the overall scale of the latent factor to prevent compensatory scaling of $F$ when adjusting hyperparameters that control the magnitude of the effect of $F$ on $t_x$ or $y$.

$$\mu_F = X_F\beta_{X_F\to F}$$
$$F \sim \sigma(\mathcal{N}(\mu_F, 1))$$
$$\beta_{X_F\to F} \sim \mathcal{N}(0, 2.5)$$

In the conditional mean function for $F$, the control variables $X_F$ were pre-centered to have zero mean.

**Proxies** We parameterize the proxies as conditionally independent Bernoulli distributed random variables. The conditional probability was implemented using the logistic link function. For each proxy $w_j$:

$$
\begin{aligned}
\eta_{w_j} &= F\beta_{F\to w_j} - \mu_{wj} \\
w_j &\sim \text{Bern}(\sigma(\eta_{wj})) \\
\beta_{F\to w_j} &\sim \text{HN}(2.5) \\
\mu_{w_j} &\sim \mathcal{N}(0, 2.5)
\end{aligned}
$$

The prior $HN(2.5)$ implements the assumption that a higher value of latent fitness leads on average to better values for the proxy variables of fitness.

**Treatment** The treatment indicator was modeled as a Bernoulli distributed random variable, with a linear model and the logistic link function, similar to the proxy variables.

$$
\begin{aligned}
\eta_{t_x} &= F\beta_{F\to t_x} + X_{t_x}\beta_{X_{t_x}\to t_x} - \mu_{t_x} \\
t_x &\sim \text{Bern}(\sigma(\eta_{t_x})) \\
\beta_{F\to t_x} &\sim \text{HN}(\sigma_{F\to t_x}) \\
\mu_{t_x} &\sim \mathcal{N}(0, 2.5)
\end{aligned}
$$

$X_{t_x}$ are all observed variables that are direct parents of treatment in the ADMG, except for $F$. $\sigma_{F\to t_x}$ is a hyperparameter that was determined per the model selection method in subsection A.1.5.

**Structural Causal Model** The entire generative model can now be formalized in a single structural causal model. Let $X^{(\cdot)}$ denote a $Nd^{(\cdot)}$ design matrix for $N$ patients with $d^{(\cdot)}$ control variables for different control variable sets, $\beta_{(.)}$ global parameters, APGW(.) the 4-parameter Adapted Power Generalized Weibull distribution [4], $w_j$ binary proxies, $t_x$ treatment variable, $y$ the survival outcome consisting of positive real number indicating time, and a binary indicator for deceased or censored.

$$
\begin{aligned}
\mu_F &= X^F\beta_{X^F\to F} \\
F &\sim \sigma(\text{N}(\mu_F, 1)) \\
\eta_{w_j} &= F\beta_{F\to w_j} - \mu_{w_j} \\
w_j &\sim \text{Bern}(\sigma(\eta_{w_j})) \\
\eta_{t_x} &= F\beta_{F\to t_x} + X_{t_x}\beta_{X_{t_x}\to t_x} - \mu_{t_x} \\
t_x &\sim \text{Bern}(\sigma(\eta_{t_x})) \\
\beta_y &= F\beta_{F\to y} + X_y\beta_{X_y\to y} + t_x(\beta_{t_x\to y} + F\beta_{F*t_x\to y} + X\beta_{X*t_x\to y}) + \beta_0 \\
y &\sim \text{APGW}(0, \beta_y, \alpha_0, \nu_0)
\end{aligned}
$$

**Effect Modifiers** Predicting treatment effects for new patients requires the estimation of the conditional average treatment effect (CATE). In the case of a binary treatment variable $t_x$ and covariates $x, w$, the CATE is defined as:

$$\text{CATE}(x, w) = \mathbb{E}\left[y|\text{do}(t_x = 1), x = x, w = w\right] - \mathbb{E}\left[y|\text{do}(t_x = 0), x = x, w = w\right] \tag{20}$$

To evaluate whether some variables are effect modifiers (i.e. the treatment effect differs between different values of this variable), we employ the definition of a conditional effect modifier by VanderWeele [20]. For covariates $x, w$ and treatment $t_x$, $w$ is said to be an effect modifier of $t_x$ conditional on $x$ if for some value of $x$ there exist two values $w_1 \neq w_2$ of $w$ for which $\text{CATE}(x, w_1) \neq \text{CATE}(x, w_2)$. Note that the estimation of a treatment effect modifier is dependent on the scale on which the CATE is measured. For a linear log-hazard ratio model (e.g. the Cox proportional hazards model or the proportional hazards APGW), this reduces to the familiar statistical interaction term $\beta_{w*t_x}$:

$$\beta(t_x, x, w) = \beta_{t_x} t_x + \beta_x x + \beta_w w + \beta_{w*t_x} w t_x \tag{21}$$

Filling in equation 21 in 20 will lead to the familiar result that the conditional average treatment effect is the average treatment effect plus $w\beta_{w*t_x}$. This treatment interaction term quantifies the difference in treatment effects between $w = 0$ and $w = 1$. Due to the linearity of this model in 21, the effect modification of $w$ is the same for all values of $x$. In our log-hazard model formulation, effect modification is similarly modeled by adding product terms between treatment and control variables, but also between treatment and the latent factor $F$. Since the conditional distribution of $p(F|X, W)$ is non-linear in $X$ and $W$, effect modification by proxies $W$ and control variables $X$ can no longer be equated to the interaction terms in the linear log-hazard model, and the effect modification of some variable $x_i$ may depend on the value of other control variables and proxy variables.

We can estimate potential effect modification by plugging our log-hazard model $\beta(t_x, F, X_y)$ defined in 18 in equation 20:

$$\text{CATE}(X, W) = \underset{F \sim p(F|X,W)}{\mathbb{E}}\left[\beta(t_x = 1, X, F)\right] - \underset{F \sim p(F|X,W)}{\mathbb{E}}\left[\beta(t_x = 0, X, F)\right] \tag{22}$$

$$= \underset{F \sim p(F|X,W)}{\mathbb{E}}\left[\beta(t_x = 1, X, F) - \beta(t_x = 0, X, F)\right] \tag{23}$$

This will estimate the log-hazard ratio between concurrent and sequential treatment as a function of $X$ and $W$. We have omitted the decency of these quantities on the global parameters $\theta$ here. As said before, the effect modification of a variable can depend on the value of the other variables due to the non-linearities in our model. To summarize the average effect modification for a unit change in a variable we use partial dependence functions [8]. Specifically we look at the average change in CATE for a unit difference in a variable $x_j$, averaged over the marginal distribution of other variables $\mathbf{x}_{-j}$.

$$\text{EM}(x_j) := \mathbb{E}_{\mathbf{x}_{-j} \sim p(\mathbf{x}_{-j})} \left[\text{CATE}(x_j = 1, \mathbf{x}_{-j}) - \text{CATE}(x_j = 0, \mathbf{x}_{-j})\right] \tag{24}$$

Where the expectation is taken over the observed data.

##### A.2.3 Model selection

As the treatment variable and the outcome variable follow different distributions there is no way to express the parameters of these models on a single scale. Specifically, the log odds ratio of fitness to treatment ($\beta_{F \to t_x}$) is incommensurable with the log hazard ratio of fitness to survival ($\beta_{F \to y}$). This creates a problem with specifying the right scale for the prior distributions of these two parameters. If the scale of the priors for one of the parameters is greater than the other, parameters that maximize the joint likelihoood can be biased towards modeling the variable with the prior with the greater scale. Therefore, the prior standard deviations on the parameters from $F$ to $y$ and from $F$ to $t_x$ were treated as hyperparameters. Values for the prior standard deviations were $\{0.1, 1.0, 2.5, 10.0, 100.0\}$, resulting in a grid of 125 hyperparameter combinations. We used 5-fold cross validation to select hyperparameters that were not refuted by the assumptions in the DAG as outlined in A.1.5. In a final inference step we follow a Bayesian Model Averaging approach by formulating a mixture model over all acceptable hyperparameter settings $\psi_j$.

$$p_{\text{BMA}}(\theta) = \sum_j p_{\psi_j}(\theta|D_{\text{full}})p(\psi_j|D_{\text{full}}) \tag{25}$$

Where each $\psi_j$ corresponds to an acceptable setting of the hyperparameters. This mixture model was evaluated once on the full dataset where global parameters and model weights were inferred jointly, resulting in our final model estimate.

##### A.2.4 Implementation

All probabilistic models were implemented using the open source probabilistic programming language NumPyro [17], version 0.4.1 with JAX [3] version 0.2.7 as a back-end. We used the No-U-Turn Hamiltonian Monte Carlo sampling algorithm to simulate 7500 samples per chain (following 2500 warm-up samples) in 4 independent chains from the posterior distribution for each hyperparameter setting. For posterior predictive densities on held-out data we numerically integrated the densities with respect to $F$ by using a fixed grid of points for $\hat{F}_\epsilon$, the noise term of $\hat{F}$. Between $-5.0$ and 5.0, 250 equally spaced values for $\hat{F}_\epsilon$ were used. From the original samples over global parameters, 1500 samples equally divided over the chains were used. The samples of global parameters were selected by slicing the original chain to reduce auto-correlation between the samples. After applying the model selection procedure we estimated the final model using 12 independent chains with 7500 samples per chain (following 2500 warm-up samples). Model evaluation was performed in R, version 4.0.3. The code that implements the statistical models will be made freely accessible online. Due to privacy regulations the clinical data cannot be made available.

Figure 5: Causal Directed Acyclic Graph for sensitivity analysis of omitted confounder. A simplified DAG is used to present the sensitivity analysis.

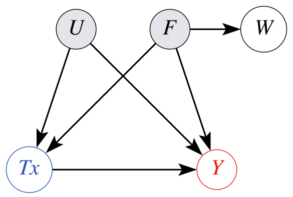

### B Sensitivity Analyses

#### B.1 Omitted Confounder

##### B.1.1 Methods

We performed a sensitivity analysis to asses the robustness of our inferences with respect to a potential unobserved confounder. Specifically, we test the effect of an unobserved confounder $U$ on the point estimate of the average treatment effect ($ATE$). The latent factor $U$ is assumed to be a direct cause of both treatment and survival, independent of the latent factor fitness, see Figure B.1.1.

Assuming a true data generating mechanism (under the sensitivity parameters $\gamma_{U\to y}, \gamma_{U\to t_x}$):

$$\begin{aligned}\beta_y = \beta_0 &+ F\beta_{F\to y} + X_y\beta_{X_y\to y} \\ &+ t_x(\beta_{t_x\to y} + F\beta_{F*t_x\to y} + X_y\beta_{X*t_x\to y}) \\ &+ \gamma_{U\to y}U\end{aligned}$$

Where $\beta_y$ is the log-hazard ratio. Whereas we estimated the treatment effect using the equivalent model for survival but omitting the term from $U$ to $y$.

As in this hypothetical setting of the sensitivity analysis we did not condition on all confounders when $|\gamma_{U\to t_x}| > 0$ and $|\gamma_{U\to y}| > 0$, the estimated treatment effect is a biased estimate of the true treatment effect. For different settings of sensitivity parameters $\gamma = \{\gamma_{U\to tx}, \gamma_{U\to y}\}$ we re-estimated the posterior distribution over global parameters, this time including $U$ as an additional latent factor with fixed coefficients to treatment and survival, as specified by $\gamma$. In order to be able to clinically reason about the likelihood of the existence of an omitted confounder $U$ that would be strong enough to alter the conclusions of our modeling, we need to be able to compare the effects of this hypothetical unobserved confounder on treatment ( $\gamma_{U\to t_x}$ ) and survival ( $\gamma_{U\to y}$ ), relative to the effects of the modeled confounder for fitness $F$ ( $\beta_{F\to t_x}, \beta_{F\to y}$ ). To make sure that the comparison of parameters was valid we fixed the standard deviation of $U$ to 1, and we re-scaled the parameters $\beta_{F\to t_x}, \beta_{F\to y}$ to the values

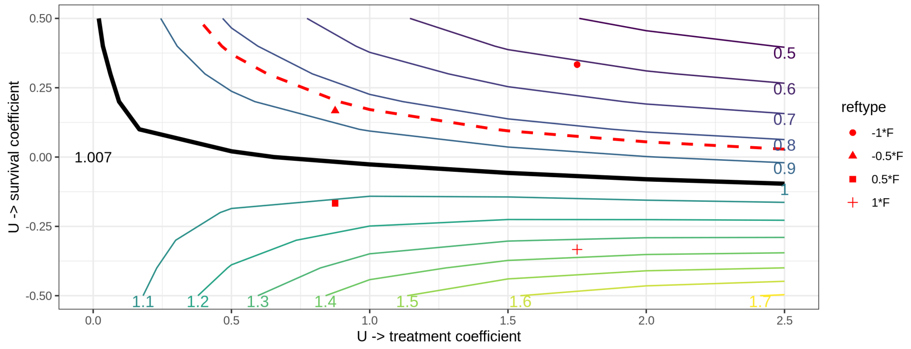

Figure 6: Results of sensitivity analysis. For different combinations of sensitivity parameters $\gamma_{U\to y}$ ($y$-axis) and $\gamma_{U\to t_x}$ ($x$-axis) we re-estimated the model, with the additional latent variable $U$. Results for the estimated average treatment effect are indicated by the level lines, marked by the point estimate of the hazard ratio for overall survival of concurrent versus sequential chemoradiation. The black line indicates the null-effect (hazard ratio $=1$). The red dotted line indicates the treatment effect reported in the meta-analysis of randomized controlled trials (hazard ratio $=0.84$) [1]. Four reference points are added to gauge how the sensitivity parameters relate to the parameters of the estimated latent confounder $F$ to treatment and survival. The reference point $1 * F$ corresponds to the value of $\hat{\beta}_{F\to y}$ and $\hat{\beta}_{F\to t_x}$ in the original model. $0.5 * F$ is the point where these coefficients are both divided by two. $-1 * F$ is the point where the sign of $\hat{\beta}_{F\to y}$ is changed, and $-0.5 * F$ is defined analogous to $0.5 * F$

they would have if $F$ were also scaled to have a standard deviation of 1. We investigate the effect of potential unobserved confounding on the treatment effect estimate for all combinations of $\gamma_{U\to y} \in \{-1, -0.5, 0, 0.5, 1\}\beta_{F\to y}$ and $\gamma_{U\to t_x} \in \{0, 0.5, 1\}\beta_{F\to t_x}$.

##### B.1.2 Results

The results of the sensitivity analysis are presented in Figure 6. If there were a confounder that was more than half as strong as the modeled latent confounder $\hat{F}$ but opposite in sign of survival, the point estimate of the $ATE$ would be greater than the estimate from the RCTs [1]. The interpretation of this latent confounder is that it increases the likelihood of being treated, but reduces the likelihood of survival. A clinical example may be that patients who have more aggressive tumors are treated more aggressively in order to improve survival outcomes.

#### B.2 Missing Weight Loss

We now describe the sensitivity analysis for the effect of different missingness mechansisms for weight loss on the estimate of the average treatment effect.

##### B.2.1 Methods

Let $M$ denote the missingness indicator for weight loss $X$, then $p(M = 1|X = x)$ denotes the probability of missingness, conditional on the value of $X$. The central assumption of this sensitivity analysis is that the missingness in weight loss is random conditional on the value of weight loss itself. Specifically, that the probability of missingness is lower when weight loss is present.

$$p(M = 1|X = 1) \leq p(M = 1|X = 0) \tag{26}$$

Applying this inequality to the marginal probability of missingness $p(M = 1)$ we get:

$$\begin{aligned} p(M = 1) &= p(M = 1|X = 1)p(X = 1) + p(M = 1|X = 0)(1 - p(X = 1)) \\ &\geq p(M = 1|X = 1)p(X = 1) + p(M = 1|X = 1)(1 - p(X = 1)) \\ &= p(M = 1|X = 1) \end{aligned}$$

It follows that $p(M = 1|X = 1) \in [0, p(M = 1)]$. From the observed data we know $p(M = 1)$, the marginal probability of missingness. We introduce sensitivity parameter $\alpha$ to parameterize the range of possible values for $p(M = 1|X = 1)$ compatible with the observed marginal probability of missingness and our assumption 26. We define:

$$p(M = 1|X = 1, \alpha) := (1 - \alpha)p(M = 1) \tag{27}$$

For $\alpha \in [0, 1]$. It then follows that $p(M = 0|X = 1, \alpha) = 1 - (1 - \alpha)p(M = 1)$. In this setup, $\alpha = 0$ represents the boundary case where the missingness in $X$ is completely at random, and $\alpha = 1$ is the boundary case where $p(M = 1|X = 1) = 0$, i.e. $X$ is always observed when $X = 1$.

Let $\pi_{\text{obs}} := p(X = 1|M = 0)$, the probability of weight loss in the observed cases and $\pi^* := p(X = 1)$ denote the unknown true marginal probability of weight loss. Using Bayes rule, we can express $\pi_{\text{obs}}$ in terms of $\pi^*$, $p(M = 1)$ and $p(M = 0|X = 1)$:

$$\begin{aligned} \pi_{\text{obs}} = p(X = 1|M = 0) &= \frac{p(X = 1)}{p(M = 0)} p(M = 0|X = 1) \\ &= \frac{\pi^*}{1 - p(M = 1)} p(M = 0|X = 1) \end{aligned}$$

Now we can write:

$$\pi^* = \frac{1 - p(M=1)}{p(M=0|X=1)} \pi_{\text{obs}} \tag{28}$$

Since we cannot estimate $p(M = 0|X = 1)$ from the observed data, we will substitute it with the $p(M = 0|X = 1, \alpha)$ using equation 27, conditional on the sensitivity parameter $\alpha$.

$$\pi^*_\alpha = \frac{1 - p(M=1)}{p(M=0|X=1,\alpha)} \pi_{\text{obs}} \tag{29}$$

Now we have that given the observed data and the sensitivity parameter $\alpha$, the unobserved quantity of interest is identified, so we can proceed with the sensitivity analysis.

**Re-weighting by weight loss** For different values of $\alpha$ we assign weights to patients based on their value for weight loss.

$$w_\alpha(x) := x w^1_\alpha + (1-x) w^0_\alpha \tag{30}$$

By assigning patients with weight loss a higher weight than patients without weight loss, we can artificially create a population with a greater prevalence of weight loss. Specifically, for each $\alpha$ we define weights such that the weighted sum of the observed values of weight loss is equal to $\pi^*_\alpha$.

$$\frac{1}{N} \sum_{i=1}^{N} [w_\alpha(x_i) x_i] = \pi^*_\alpha$$

By further requiring that the sum of weights is equal to the total number of patients,

$$\sum_{i=1}^{N} w_\alpha(x_i) = N$$

the weights are now uniquely defined as:

$$w^1_\alpha = \frac{\pi^*_\alpha}{\pi_{\text{obs}}}$$
$$w^0_\alpha = \frac{1 - w^1_\alpha \pi_{\text{obs}}}{1 - \pi_{\text{obs}}}$$

Using these weights, for each $\alpha$ we calculate a new average treatment effect by taking the weighted mean of the conditional treatment effects:

$$\text{ATE}(\alpha) = \frac{1}{N} \sum_{i=1}^{N} w_\alpha(x_i) \text{CATE}_i \tag{31}$$

##### B.2.2 Results

The results of this sensitivity analysis are presented in Table B.2.2 and Figure B.2.2. The ATE shows a minor shift in the direction of concurrent being more effective when the dependency of missingness on the actual value of weight loss becomes stronger. This is explained by the fact that patients with weight loss are estimated to have less benefit of concurrent treatment. If missingness were always observed if it was present, this means that the true average value of weight loss is lower in the population than when weight loss is missing completely at random.

| $\alpha$ | $p(M=1\|X=1)$ | $p(M=1\|X=0)$ | $\pi^*_\alpha$ | $w^1$ | $w^0$ | hazard ratio | CI low | CI high |
|---|---|---|---|---|---|---|---|---|
| 0.0 | 0.448 | 0.170 | 0.170 | 1.000 | 1.000 | 1.011 | 0.650 | 1.534 |
| 0.1 | 0.439 | 0.153 | 0.183 | 1.020 | 0.983 | 1.013 | 0.689 | 1.630 |
| 0.2 | 0.431 | 0.136 | 0.195 | 1.041 | 0.967 | 1.015 | 0.650 | 1.536 |
| 0.3 | 0.423 | 0.119 | 0.207 | 1.061 | 0.950 | 1.017 | 0.691 | 1.636 |
| 0.4 | 0.415 | 0.102 | 0.218 | 1.082 | 0.934 | 1.019 | 0.693 | 1.639 |
| 0.5 | 0.407 | 0.085 | 0.228 | 1.102 | 0.917 | 1.020 | 0.706 | 1.672 |
| 0.6 | 0.399 | 0.068 | 0.237 | 1.123 | 0.900 | 1.022 | 0.706 | 1.673 |
| 0.7 | 0.392 | 0.051 | 0.246 | 1.143 | 0.884 | 1.024 | 0.706 | 1.675 |
| 0.8 | 0.385 | 0.034 | 0.255 | 1.163 | 0.867 | 1.026 | 0.709 | 1.681 |
| 0.9 | 0.379 | 0.017 | 0.263 | 1.184 | 0.850 | 1.028 | 0.710 | 1.682 |
| 1.0 | 0.372 | 0.000 | 0.270 | 1.204 | 0.834 | 1.030 | 0.711 | 1.684 |

Results of sensitivity analysis to missing data in weight loss. $\alpha$: sensitivity parameter, $M$ missingness indicator, $X$ indicator for presence of weight loss, $\pi^*_\alpha$ true marginal probability of weight loss under sensitivity parameter $\alpha$, $w^1$ weight for patients with weight loss, $w^0$ weight for patients without weight loss, hazard ratio: point estimate of the average treatment effect of concurrent versus sequential chemoradiation for overall survival. CI: 95% credible interval

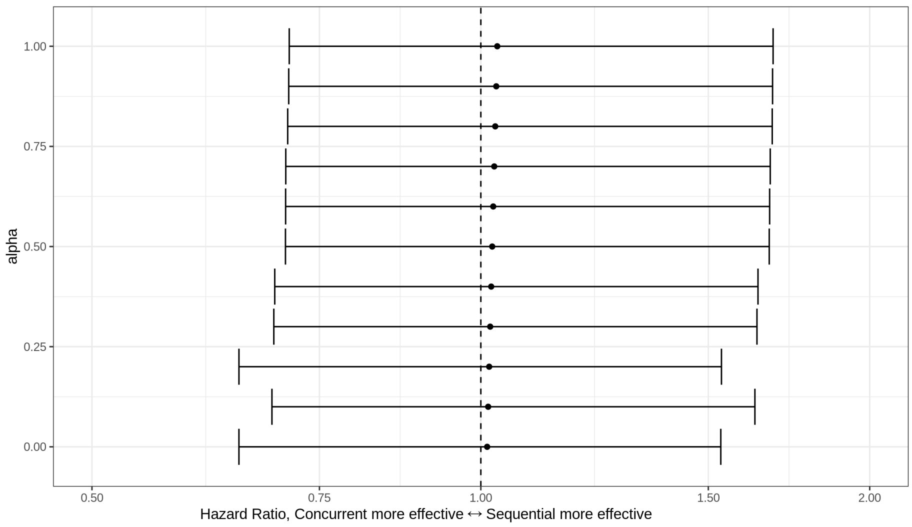

Figure 7: Different average treatment effect values for different values of sensitivity parameter $\alpha$ with 95% credible interval

### C Extrapolation to Randomized Trials

#### C.1 Methods

A central assumption in our study is that the average treatment effect reported in randomized trials is not directly transportable to our population due to differences between the patient populations. We assume however that the conditional treatment effect is transportable. This assumption implies that a patient in our population has the same benefit and harms of treatment as a similar patient in the randomized trials has. Similar here means that they have the same values for the covariates, latent tumor behavior and latent fitness. Using our estimate of the conditional treatment effect we can extrapolate our results to calculate what the estimated average treatment effect would be if our population were more alike the population from the randomized trials.

**Matching the RCT Population** Average descriptive statistics on the study population of the RCTs (e.g. the mean age) are available from [1]. However, matching our population to theirs is not possible based on these criteria alone. In addition to the published inclusion and exclusion criteria for each RCT included in [1] we should expect hidden confounding between inclusion in the trial and overall survival [7]. Patients included in the randomized trials should be able to undergo all treatment arms. This means that patients who were deemed clinically unfit for concurrent treatment can be expected to be underrepresented in the RCT population. The assessment of this fitness for inclusion in the RCT is essentially the same as the assessment of fitness for concurrent treatment in

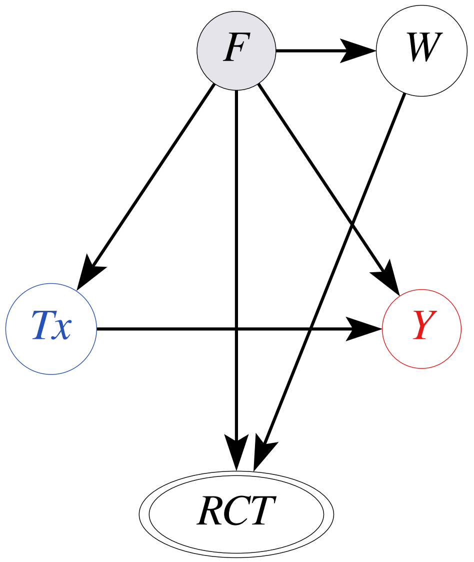

Figure 8: Directed Acyclic Graph of inclusion in RCTs. The same (unobserved) confounding that influences treatment decisions in real-world practice will also influence inclusion in the RCTs. In addition to the unobserved confounder fitness, the concrete exclusion criteria based on proxy measurements (e.g. performance score) also determine inclusion in the RCT

our real-world population, with the addition of strict inclusion criteria from the trials. See Figure 8 for a DAG that depicts this mechanism. Using the predicted probability of concurrent treatment from the model, we can restrict our population to a subpopulation with higher fitness using different cut-offs for the predicted probability of concurrent treatment.

#### C.2 Results

We found that the estimated Average Treatment Effect increased in favor of concurrent treatment when restricting the population to higher predicted probability of concurrent treatment, see Figure 9.

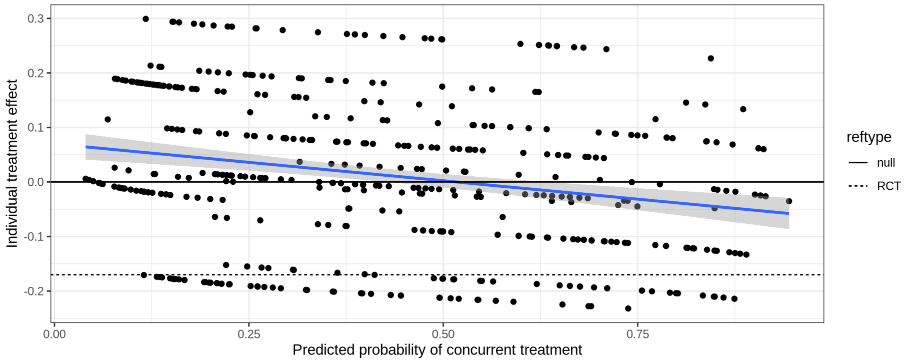

Figure 9: Predicted probability of concurrent treatment versus estimated individual treatment effect per patient. When trying to match the RCT by restricting the population to those with a higher predicted probability of concurrent treatment, the estimated average treatment effect becomes closer to that of the RCTs [1]. Added is a linear regression fit line with 95% prediction confidence band. Reference lines are added for the null-effect (hazard ratio 1) and the effect reported in the randomized trials (hazard ratio 0.84) [1].

### D Results

#### D.1 Cohort

Patients were recruited from 9 different hospitals in the Utrecht region of the Netherlands. Specifically, the University Medical Center Utrecht, the Antonius Hospital, locations Nieuwegein and Woerden, the Meander Medical Center Amersfoort, the Diakonessen Hospital, Utrecht, the Beatrix Hospital, Gorinchem, the Haaglanden Medical Center, The Hague, the Amsterdam Medical Center, Amsterdam, the Gelderse Vallei Hospital, Ede, and the Antoni van Leeuwenhoek Hospital. These hospitals represent a rich ensemble of university hospitals, specialized oncological hospitals and both large and small secondary care institutions.

All patients were referred to the University Medical Center Utrecht for radiotherapy, but their care was coordinated by the physician in the referring hospital.

Baseline data of our cohort and that of the published meta-analysis of randomized clinical trials [1] are presented in Table 1. Percentages were calculated on the available data. Note that our cohort is older, has worse performance scores and is less frequently treated with concurrent therapy.

#### D.2 Supplemental Tables

| | ours | RCT |
|---|---|---|
| n | 504 | 1205 |
| age (%) | | |
| <60 | 143 (28.4) | 519 (43.1) |
| 60-64 | 85 (16.9) | 225 (18.7) |
| 65-69 | 97 (19.2) | 270 (22.4) |
| >=70 | 179 (35.5) | 189 (15.7) |
| missing | 0 (0.0) | 2 (0.2) |
| histology (%) | | |
| adeno | 200 (39.7) | 395 (32.8) |
| squamous | 194 (38.5) | 549 (45.6) |
| other | 89 (17.7) | 256 (21.2) |
| missing | 21 (4.2) | 5 (0.4) |
| male sex (%) | 300 (59.5) | 921 (76.4) |
| stage (%) | | |
| I | 0 (0.0) | 6 (0.5) |
| II | 0 (0.0) | 12 (1.0) |
| IIIA | 268 (53.2) | 441 (36.6) |
| IIIB | 232 (46.0) | 735 (61.0) |
| IIIC | 2 (0.4) | 0 (0.0) |
| IV | 0 (0.0) | 3 (0.2) |
| missing | 2 (0.4) | 8 (0.7) |

Table 1: Baseline characteristics of participants in our study (ours) and in the meta-analysis of randomized trial (RCTs) [1]

Table 2: Statistics of MCMC samples of parameters of the final bayesian model average. The variables F_mu, F_sd and b_txbinary_y_marginal are not parameters of the model, but are deterministically calculated from the parameters, and are included for reference. The parameters pmodel[0], pmodel[1] and pmodel[2] are the model probabilities for the three selected hyperparameter settings: $\sigma_{F \to t_x} = 2.5$, $\sigma_{F \to y} \in \{0.1, 1.0, 2.5\}$. The parameters are reported on the scale of the linear model (i.e. log odds ratios for the binary proxies and treatment, and log hazard ratios for the outcome survival). Given that a higher hazard means a worse overall survival, the negative sign of b_F_y indicates that higher fitness leads to better overall survival. At the same time the positive sign of b_F_tx indicates that higher fitness leads to a higher chance of concurrent treatment

| | mean | sd | hdi_2.5% | hdi_97.5% | mcse_mean | mcse_sd |
|---|---|---|---|---|---|---|
| b_F_eGFRunder60 | -2.300 | 0.878 | -4.045 | -0.631 | 0.004 | 0.003 |
| b_F_ecogbinary1 | -2.047 | 0.575 | -3.186 | -0.940 | 0.004 | 0.003 |
| b_F_tx | 6.157 | 1.486 | 3.296 | 9.061 | 0.013 | 0.009 |
| b_F_y | -1.171 | 0.731 | -2.511 | 0.125 | 0.006 | 0.005 |
| b_Ftx_y | -0.122 | 0.354 | -0.814 | 0.572 | 0.001 | 0.001 |
| b_agectd_F | -0.873 | 0.270 | -1.394 | -0.466 | 0.003 | 0.002 |
| b_histoother_F | -0.189 | 0.573 | -1.338 | 0.938 | 0.003 | 0.002 |
| b_histoother_tx | 0.113 | 0.664 | -1.211 | 1.425 | 0.003 | 0.002 |
| b_histoother_y | 0.013 | 0.210 | -0.401 | 0.429 | 0.001 | 0.001 |
| b_histoothertx_y | -0.085 | 0.164 | -0.409 | 0.236 | 0.000 | 0.000 |
| b_histosquamous_F | -0.878 | 0.527 | -1.964 | 0.102 | 0.003 | 0.002 |
| b_histosquamous_tx | 0.089 | 0.618 | -1.084 | 1.338 | 0.003 | 0.002 |
| b_histosquamous_y | 0.226 | 0.198 | -0.170 | 0.599 | 0.001 | 0.001 |
| b_histosquamoustx_y | -0.114 | 0.156 | -0.420 | 0.190 | 0.000 | 0.000 |
| b_sIIIB_F | -0.517 | 0.492 | -1.495 | 0.421 | 0.002 | 0.002 |
| b_sIIIB_tx | -0.771 | 0.545 | -1.841 | 0.331 | 0.002 | 0.002 |
| b_sIIIB_y | 0.133 | 0.157 | -0.176 | 0.443 | 0.001 | 0.000 |
| b_sIIIBtx_y | 0.169 | 0.151 | -0.126 | 0.465 | 0.000 | 0.000 |
| b_txbinary_y | -0.019 | 0.322 | -0.645 | 0.614 | 0.002 | 0.001 |
| b_wtlossany_F | -1.273 | 0.566 | -2.395 | -0.255 | 0.004 | 0.003 |
| b_wtlossany_tx | 1.494 | 0.650 | 0.244 | 2.778 | 0.003 | 0.002 |
| b_wtlossany_y | -0.135 | 0.202 | -0.539 | 0.246 | 0.001 | 0.001 |
| b_wtlossanytx_y | 0.161 | 0.155 | -0.140 | 0.467 | 0.000 | 0.000 |
| mu_eGFRunder60 | 1.251 | 0.376 | 0.508 | 1.981 | 0.002 | 0.001 |
| mu_ecogbinary1 | -0.838 | 0.293 | -1.418 | -0.273 | 0.002 | 0.001 |
| mu_tx | 3.667 | 1.051 | 1.698 | 5.775 | 0.008 | 0.006 |
| alpha0 | 0.743 | 0.069 | 0.608 | 0.880 | 0.000 | 0.000 |
| beta0 | -0.276 | 0.382 | -1.010 | 0.467 | 0.003 | 0.002 |
| nu0 | -0.061 | 0.057 | -0.169 | 0.054 | 0.000 | 0.000 |
| F_mu | 0.482 | 0.017 | 0.449 | 0.514 | 0.000 | 0.000 |
| F_sd | 0.285 | 0.026 | 0.242 | 0.337 | 0.000 | 0.000 |
| b_txbinary_y_marginal | 0.010 | 0.223 | -0.409 | 0.450 | 0.002 | 0.001 |
| pmodel[0] | 0.272 | 0.209 | 0.000 | 0.682 | 0.001 | 0.000 |
| pmodel[1] | 0.371 | 0.242 | 0.000 | 0.808 | 0.001 | 0.000 |
| pmodel[2] | 0.358 | 0.241 | 0.000 | 0.797 | 0.001 | 0.001 |

Table 3: Sampling statistics for parameters of the final bayesian model average. The variables F_mu, F_sd and b_txbinary_y_marginal are not parameters of the model, but are deterministically calculated from the parameters, and are included for reference. The parameters pmodel[0], pmodel[1] and pmodel[2] are the model probabilities for the three selected hyperparameter settings: $\sigma_{F\to t_x} = 2.5$, $\sigma_{F\to y} \in \{0.1, 1.0, 2.5\}$

| | ess_mean | ess_sd | ess_bulk | ess_tail | r_hat |
|---|---|---|---|---|---|
| b_F_eGFRunder60 | 50491.0 | 50491.0 | 47940.0 | 57108.0 | 1.0 |
| b_F_ecogbinary1 | 20092.0 | 20092.0 | 19587.0 | 31950.0 | 1.0 |
| b_F_tx | 12583.0 | 12583.0 | 12328.0 | 21622.0 | 1.0 |
| b_F_y | 13274.0 | 12712.0 | 14190.0 | 21844.0 | 1.0 |
| b_Ftx_y | 112564.0 | 79754.0 | 112564.0 | 97384.0 | 1.0 |
| b_agectd_F | 11479.0 | 11479.0 | 12611.0 | 19832.0 | 1.0 |
| b_histoother_F | 50821.0 | 35482.0 | 53186.0 | 52046.0 | 1.0 |
| b_histoother_tx | 63335.0 | 63335.0 | 63735.0 | 75460.0 | 1.0 |
| b_histoother_y | 80292.0 | 62519.0 | 81810.0 | 77530.0 | 1.0 |
| b_histoothertx_y | 211765.0 | 73591.0 | 211766.0 | 95551.0 | 1.0 |
| b_histosquamous_F | 38386.0 | 30373.0 | 41861.0 | 41783.0 | 1.0 |
| b_histosquamous_tx | 57878.0 | 57878.0 | 59599.0 | 68582.0 | 1.0 |
| b_histosquamous_y | 41188.0 | 41188.0 | 43284.0 | 54958.0 | 1.0 |
| b_histosquamoustx_y | 196417.0 | 85781.0 | 196450.0 | 94168.0 | 1.0 |
| b_sIIIB_F | 42614.0 | 24811.0 | 49835.0 | 41523.0 | 1.0 |
| b_sIIIB_tx | 64364.0 | 64364.0 | 65085.0 | 73268.0 | 1.0 |
| b_sIIIB_y | 93295.0 | 90465.0 | 95162.0 | 76567.0 | 1.0 |
| b_sIIIBtx_y | 223477.0 | 115619.0 | 223486.0 | 92992.0 | 1.0 |
| b_txbinary_y | 29011.0 | 29011.0 | 30389.0 | 37142.0 | 1.0 |
| b_wtlossany_F | 26035.0 | 24869.0 | 28244.0 | 37178.0 | 1.0 |
| b_wtlossany_tx | 46061.0 | 46061.0 | 44526.0 | 49248.0 | 1.0 |
| b_wtlossany_y | 32376.0 | 32376.0 | 33430.0 | 50794.0 | 1.0 |
| b_wtlossanytx_y | 199570.0 | 107291.0 | 199624.0 | 94483.0 | 1.0 |
| mu_eGFRunder60 | 47445.0 | 45579.0 | 47018.0 | 61435.0 | 1.0 |
| mu_ecogbinary1 | 22216.0 | 22216.0 | 21742.0 | 36303.0 | 1.0 |
| mu_tx | 16716.0 | 16716.0 | 16307.0 | 28481.0 | 1.0 |
| alpha0 | 188062.0 | 188062.0 | 190422.0 | 94694.0 | 1.0 |
| beta0 | 17370.0 | 17370.0 | 17917.0 | 29165.0 | 1.0 |
| nu0 | 75598.0 | 75598.0 | 78518.0 | 66719.0 | 1.0 |
| F_mu | 87959.0 | 87959.0 | 87552.0 | 85513.0 | 1.0 |
| F_sd | 12815.0 | 12677.0 | 13582.0 | 21807.0 | 1.0 |
| b_txbinary_y_marginal | 15367.0 | 15367.0 | 16531.0 | 23319.0 | 1.0 |
| pmodel[0] | 144220.0 | 105733.0 | 143474.0 | 67157.0 | 1.0 |
| pmodel[1] | 171243.0 | 134171.0 | 165659.0 | 73870.0 | 1.0 |
| pmodel[2] | 137870.0 | 110311.0 | 134905.0 | 72606.0 | 1.0 |

### E Discussion

#### E.1 Potential Applications of PROTECT

We now provide two examples where the PROTECT method may be used.

**example 1: laryngeal carcinoma** For locally advanced unresectable squamous cell laryngeal carcinoma, concurrent chemoradiation is recommended. For patients over 70 years of age or who are unfit for concurrent treatment, only radiotherapy is recommended [12]. RCTs comparing concurrent chemoradiation with radiotherapy alone will be conducted in populations where concurrent chemoradiation is a feasible treatment. Therefore, the RCTs provide no evidence on the treatment effect of concurrent treatment for older and weaker patients. In real-world clinical practice, older and weaker patients may receive the concurrent treatment for example when they have a strong preference for this treatment. Observational studies comparing both treatments will have to deal with the same unobserved confounder as in the NSCLC case: the overall fitness of the patient. To estimate the treatment effect in the older and weaker population, PROTECT can be used to estimate the treatment effect from observational data.

**example 2: esophageal cancer** The second example concerns stage III squamous cell esophageal cancer. For these patients, neoadjuvant chemoradiation followed by esophagectomy is considered the most effective treatment while definitive chemoradiation without surgery is recommended for patients who are unfit for surgery or refuse surgery [10]. Whether chemoradiation followed by surgery should be preferred for every patient who is fit enough for surgery remains unanswered, as all RCTs are conducted with only patients who are fit for surgery and receive some form of surgical treatment [9, 5], or with patients who are unfit for surgery and never receive surgical treatment [13, 6]. In real-world clinical practice there will be patients who are deemed fit enough for surgery but refuse the surgery due to reasons not related to their prognosis, e.g. personal preference. Retrospective comparisons of chemoradiation with surgery versus chemoradiation alone will arguably be biased due to the same unobserved confounding as in the case of stage III NSCLC: the overall fitness of the patient, this time specifically fitness for surgical treatment. To answer the question whether surgery should be preferred for every patient who is fit enough, the PROTECT method could be applied to to observational data to mitigate the confounding bias.

#### References

[1] Anne Aupérin et al. "Meta-analysis of concomitant versus sequential radiochemotherapy in locally advanced non-small-cell lung cancer". en. In: *J. Clin. Oncol.* 28.13 (May 2010), pp. 2181–2190.

[2] Kenneth A. Bollen. "Confirmatory Factor Analysis". en. In: *Structural Equations with Latent Variables.* Section: Seven _eprint: https://onlinelibrary.wiley.com/doi/pdf/10.1002/97811 John Wiley & Sons, Ltd, 1989, pp. 226–318. ISBN: 978-1-118-61917-9. DOI: `10.1002/9781118619179.ch7`. URL: `https://onlinelibrary.wiley.com/doi/abs/10.1002/9781118619179.ch7` (visited on 12/28/2020).

[3] James Bradbury et al. *JAX: composable transformations of Python+NumPy programs.* 2018. URL: `http://github.com/google/jax`.

[4] Kevin Burke, M. C. Jones, and Angela Noufaily. "A Flexible Parametric Modelling Framework for Survival Analysis". en. In: *Journal of the Royal Statistical Society: Series C (Applied Statistics)* 69.2 (Apr. 2020). Number: 2, pp. 429–457. ISSN: 1467-9876. URL: `http://oro.open.ac.uk/69193/` (visited on 01/11/2021).

[5] Federico Coccolini et al. "Neoadjuvant chemotherapy in advanced gastric and esophago-gastric cancer. Meta-analysis of randomized trials". eng. In: *International Journal of Surgery (London, England)* 51 (Mar. 2018), pp. 120–127. ISSN: 1743-9159. DOI: `10.1016/j.ijsu.2018.01.008`.

[6] Thierry Conroy et al. "Definitive chemoradiotherapy with FOLFOX versus fluorouracil and cisplatin in patients with oesophageal cancer (PRODIGE5/ACCORD17): final results of a randomised, phase 2/3 trial". eng. In: *The Lancet. Oncology* 15.3 (Mar. 2014), pp. 305–314. ISSN: 1474-5488. DOI: `10.1016/S1470-2045(14)70028-2`.

[7] Issa J. Dahabreh, James M. Robins, and Miguel A. Hernán. "Benchmarking Observational Methods by Comparing Randomized Trials and Their Emulations". eng. In: *Epidemiology (Cambridge, Mass.)* 31.5 (Sept. 2020), pp. 614–619. ISSN: 1531-5487. DOI: `10.1097/EDE.0000000000001231`.

[8] Jerome H. Friedman. "Greedy function approximation: A gradient boosting machine." en. In: *Annals of Statistics* 29.5 (Oct. 2001). Publisher: Institute of Mathematical Statistics, pp. 1189–1232. ISSN: 0090-5364, 2168-8966. DOI: `10.1214/aos/1013203451`. URL: `https://projecteuclid.org/euclid.aos/1013203451` (visited on 12/31/2020).

[9] P. van Hagen et al. "Preoperative chemoradiotherapy for esophageal or junctional cancer". eng. In: *The New England Journal of Medicine* 366.22 (May 2012), pp. 2074–2084. ISSN: 1533-4406. DOI: `10.1056/NEJMoa1112088`.

[10] A. Ajani Jaffer. *NCCN Esophageal and Esophagogastric Junction Cancers, Version 1.2021.* Feb. 2021. URL: `https://www.nccn.org/professionals/physician_gls/pdf/esophageal.pdf` (visited on 02/15/2021).

[11] Christos Louizos et al. "Causal Effect Inference with Deep Latent-Variable Models". In: (May 2017). arXiv: `1705.08821 [stat.ML]`.

[12] J.-P. Machiels et al. "Squamous cell carcinoma of the oral cavity, larynx, oropharynx and hypopharynx: EHNS–ESMO–ESTRO Clinical Practice Guidelines for diagnosis, treatment and follow-up†". English. In: *Annals of Oncology* 31.11 (Nov. 2020). Publisher: Elsevier, pp. 1462–1475. ISSN: 0923-7534, 1569-8041. DOI: 10.1016/j.annonc.2020.07.011. URL: https://www.annalsofoncology.org/article/S0923-7534(20)39949-X/abstract (visited on 02/23/2021).

[13] Bruce D. Minsky et al. "INT 0123 (Radiation Therapy Oncology Group 94-05) phase III trial of combined-modality therapy for esophageal cancer: high-dose versus standard-dose radiation therapy". eng. In: *Journal of Clinical Oncology: Official Journal of the American Society of Clinical Oncology* 20.5 (Mar. 2002), pp. 1167–1174. ISSN: 0732-183X. DOI: 10.1200/JCO.2002.20.5.1167.

[14] M M Oken et al. "Toxicity and response criteria of the Eastern Cooperative Oncology Group". en. In: *Am. J. Clin. Oncol.* 5.6 (Dec. 1982), pp. 649–655.

[15] "Causal Diagrams and the Identification of Causal Effects". In: *Causality*. Ed. by Judea Pearl. Cambridge: Cambridge University Press, 2009, pp. 79–80. ISBN: 978-0-521-89560-6. DOI: 10.1017/CBO9780511803161.005. URL: https://www.cambridge.org/core/books/causality/causal-diagrams-and-the-identification-of-causal-effects/D9AE074727C3AC9AFE9F0CD4C7A506B5 (visited on 12/28/2020).

[16] Judea Pearl. *Causality*. en. Cambridge University Press, Sept. 2009.

[17] Du Phan, Neeraj Pradhan, and Martin Jankowiak. "Composable Effects for Flexible and Accelerated Probabilistic Programming in NumPyro". In: *arXiv preprint arXiv:1912.11554* (2019).

[18] Ricardo Silva and Zoubin Ghahramani. "The Hidden Life of Latent Variables: Bayesian Learning with Mixed Graph Models". en. In: (), p. 52.

[19] Anders Skrondal and Sophia Rabe-Hesketh. "5.2.5 Empirical identification". en. In: *Generalized Latent Variable Modeling: Multilevel, Longitudinal, and Structural Equation Models*. URL: https://www.routledge.com/Generalized-Latent-Variable-Modeling-Multilevel-Longitudinal-and-Structural/Skrondal-Rabe-Hesketh/p/book/9781584880004 (visited on 03/03/2021).

[20] Tyler J. VanderWeele. "On the distinction between interaction and effect modification". eng. In: *Epidemiology (Cambridge, Mass.)* 20.6 (Nov. 2009), pp. 863–871. ISSN: 1531-5487. DOI: 10.1097/EDE.0b013e3181ba333c.

[21] Aki Vehtari, Andrew Gelman, and Jonah Gabry. "Practical Bayesian model evaluation using leave-one-out cross-validation and WAIC". en. In: *Statistics and Computing* 27.5 (Sept. 2017), pp. 1413–1432. ISSN: 0960-3174, 1573-1375. DOI: 10.1007/s11222-016-9696-4. URL: http://link.springer.com/10.1007/s11222-016-9696-4 (visited on 10/05/2020).

[22] Ian R. White and John B. Carlin. "Bias and efficiency of multiple imputation compared with complete-case analysis for missing covariate values". en. In: *Statistics in Medicine* 29.28 (2010). _eprint: https://onlinelibrary.wiley.com/doi/pdf/10.1002/sim.3944, pp. 2920–2931. ISSN: 1097-0258. DOI: 10.1002/sim.3944. URL: https://onlinelibrary.wiley.com/doi/abs/10.1002/sim.3944 (visited on 06/30/2020).
